# Multivariate Prediction of Conductive Dysfunction in Well and NICU Newborns using Wideband Acoustic Immittance with Acoustic Reflex Tests

**DOI:** 10.64898/2026.03.13.26348314

**Authors:** Lisa L. Hunter, M. Patrick Feeney, Denis F. Fitzpatrick, Douglas H. Keefe

## Abstract

**Objectives:** The overall goal of this study was to assess tympanometric and ambient wideband acoustic immittance (WAI) tests and wideband acoustic reflex thresholds (ART) in well-baby and newborn intensive care (NICU) cohorts with three specific objectives: 1) Assess predictive accuracy for WBT and ART for conductive dysfunction in ears referring on the first or second stages of newborn hearing screening; 2) Identify inadequate tests likely due to probe blockages or leaks; and 3) Assess prediction models separately for well-baby and NICU screening outcomes.

**Design:** Prospective, observational study of full-term (n=514) and premature newborns (n=239) recruited from well-baby and NICU nursery birth hospital newborn hearing screening program. Wideband tympanometry, ambient absorbance, and acoustic reflexes were tested after Stage 1 transient otoacoustic emissions (TEOAE) screening. The reference standard for Pass or Refer groups was initially defined on the stage 1 TEOAE test result. Pass or Refer groups were then reassigned based on the stage 2 screening ABR for those who referred at Stage 1, and all NICU infants. Multivariate models were developed using reflectance and admittance variables to predict conductive dysfunction relative to the screening reference standard in a randomized sub-group of subjects at Stage 1 and Stage 2 screening. Classification accuracy was evaluated on a second, independent sub-group. Individual tests were classified as having inadequate probe fits if they had excessively low values of sound pressure level or susceptance (leak) or absorbance (blockage).

**Results:** Differences in ambient absorbance for Pass v. Refer screening groups revealed the greatest differences and effect sizes occurring in frequency bins between 1.4-2 kHz. Screening failure at both Stage 1 and 2 was most accurately predicted by models using ambient absorbance and power level variables at frequencies between 1-2.8 kHz, including ARTs. Tympanometric admittance variables at the positive-pressure tail for frequencies between 1-2.8 kHz in combination with the ART were more accurate predictors than those at peak pressure or the negative-pressure tail. Multivariate models generalized well to an independent group of infants at both Stage 1 and 2 for both the ambient and tympanometric models. Ambient tests revealed more inadequate tests than tympanometric tests, primarily due to blocked probe tips. Exclusion of ears to detect probe leaks or blockages slightly improved the ambient prediction models, but did not affect tympanometric models.

**Conclusion:** Wideband acoustic reflex tests improved all models for ambient and tympanometric absorbance. Multivariate prediction models developed for WAI tests were repeatable in an independent group of well and NICU infants, suggesting that the results are generalizable to these populations. Detection of probe blockage or leaks slightly improved prediction for ambient measures. Pressurized tests have the advantage of ensuring probe seals due to the need for a hermetic seal, thus are useful to ensure adequate probe insertion.

## I. INTRODUCTION

The goal of newborn hearing screening (NHS) is to detect permanent congenital hearing loss, but high rates of temporary middle-ear fluid and debris in the ear canal result in low positive predictive value, even after repeated screening. While about 3% of infants refer after two screens, only 5-10% of these cases are later diagnosed with permanent hearing loss (Centers for Disease & Prevention, 2003). Temporary, conductive hearing loss can be prolonged in the first year of life (Doyle et al., 2000). Middle-ear fluid present at birth often results in screening failure, requires repeat screening and diagnostic testing to determine if the loss is permanent, can complicate or delay diagnosis, and if persistent, requires medical management (Boudewyns et al., 2011; Doyle et al., 2000; Rosenfeld et al., 2016).

Debris (e.g. vernix) in the outer ear canal can occlude the probe tip and result in newborn hearing screening failure, while immature compliance of the newborn ear canal can cause physical occlusion (Doyle et al., 2000; Maxon et al., 1997). These conditions are difficult to detect in newborns and are considered “false positive screens.” However, true cases of permanent sensorineural or conductive hearing loss can be missed if the screening result is falsely assumed to be due to temporary middle ear fluid.

Two-stage hearing screening is common, e.g., either repeated otoacoustic emissions, or a second test with automated auditory brainstem response (ABR) to increase specificity and lower false-positive rates (Porter et al., 2009). A clinically accepted gold standard middle-ear test at the time of birth screenings does not currently exist. The accepted standard for diagnosing conductive hearing loss following repeated screen failure is diagnostic, frequency-specific threshold ABR with bone conduction (JCIH 2019). Detecting transient conductive dysfunction at the time of initial screening could streamline protocols by identifying infants most likely to have transient versus permanent congenital hearing loss, who do not require repeated screening before diagnostic audiology referral (Hunter et al., 2010). Infants with persistent middle ear dysfunction could then be referred to otolaryngology for appropriate assessment and treatment, and followed up by audiology to exclude underlying permanent hearing loss.

### 1.1. Wideband acoustic immittance (WAI) tests

Wideband acoustic immittance (WAI) tests have been developed and applied to improve detection of middle-ear dysfunction in newborns and infants (Keefe et al., 1993). WAI is a family of middle-ear acoustic measures that use click stimuli, analyzed across a broad frequency range (0.2 to 8 kHz) usually expressed as reflectance or absorbance, but WAI may also be analyzed as impedance or admittance variables (Allen et al., 2005; Keefe et al., 2000). Ambient-pressure wideband (WB) reflectance (Allen et al., 2005; Keefe et al., 2000; Keefe, Gorga, et al., 2003; Keefe, Zhao, et al., 2003) and pressurized WB tympanometry (Sanford et al., 2009b) have shown higher prediction of distortion product otoacoustic emissions (DPOAE) newborn screening test status. Area under the receiver operating characteristic curve (AUC) of 0.78 to 0.90 has been reported, compared to 0.53 to 0.75 for 1-kHz tympanometry (Aithal et al., 2015; Hunter et al., 2010; Sanford et al., 2009a). A gap in previous research concerns the issue that NICU infants have more immature middle ears and are at greater risk of both conductive and SNHL (Hunter et al., 2016; Shahnaz, 2008). This higher-risk group has not previously been compared to well babies on WAI test performance, and results could be different than in well babies.

Wideband acoustic reflex tests (WB ART) have substantially lower reflex thresholds, so that stimulus levels are safer and better tolerated in infants (Feeney & Keefe, 1999; Feeney & Sanford, 2005). The WB reflex is absent in neural causes of hearing loss, important for high-risk infants cared for in the NICU (JCIH, 2019). The ipsilateral test mode allows use of a single probe, compared to contralateral reflex measurement (Keefe et al., 2010). The first report of ambient pressure WB ART in infants showed that reflex thresholds were elevated by 24 dB in NHS refer ears compared to pass ears (Keefe et al., 2010). A longitudinal study of pressurized WB ART from birth to 12 months in well babies and babies cared for in newborn intensive care units (NICUs) showed significantly higher WB ART thresholds in ears with conductive or sensorineural hearing loss and lower thresholds at tympanometric peak pressure (TPP) (Hunter et al., 2017). The combination of WB tympanometry and WB ART has not previously been reported in infants screened in an NHS program.

### 1.2. Development of prediction models for WAI

Univariate and multivariate analyses of WAI tests have been used to identify neonatal ears at risk for conductive hearing loss, in which a model with the best performance based on AUC is constructed to classify each ear as pass or fail (Aithal et al., 2015; Hunter et al., 2010; Keefe, Gorga, et al., 2003; Sanford et al., 2009b). While multivariate models have reported higher AUC values than univariate models, there is a risk of over-fitting the model, which may generalize poorly to a different sample, so a smaller number of variables can be selected using principle components analysis (PCA) (Keefe, Gorga, et al., 2003; Keefe, Zhao, et al., 2003). Development of a multivariate model in one group of ears with validation in a second sample was reported by Myers et al. (2018), limiting the WB predictor variables to those with the highest univariate AUC values in addition to variables previously reported to be important (e.g., at 1, 2 and 4 kHz). The resulting AUCs were similar in development (0.88) and validation (0.90) groups (Myers et al., 2018).

### 1.3. Generalizability of WB Tests

Three key factors that may inflate AUC values are also important to control. First, Myers et al. (2018) selected referrals based on both 1 kHz tympanometry and DPOAE, excluding ears that passed only one of these tests. This method does not represent typical newborn screening referrals, due to the exclusion of ears with ambiguous results. Second, the validation group was composed of the opposite test ear of infants included in the development group. WB tests in left and right ears are more highly correlated in the same person than across different participants (Werner et al., 2010), which may inflate the agreement. To address this issue, both ears from each infant can be assigned to the same development or validation group (Keefe, Gorga, et al., 2003). Lastly, WB variables such as absorbance across different frequencies are correlated, violating the principle of statistical independence (Werner et al., 2010). The present study used four procedures to develop clinically generalizable multivariate prediction models: 1. NHS failure was based on the standard two-stage process of TEOAE and ABR screening; 2. The training and validation groups kept both ears of an individual together to avoid inflating prediction; 3. AUC was evaluated with and without probe fit exclusions; 4. The overall measure of prediction performance was evaluated by the AUC in the evaluation group for well and NICU cases.

### 1.4. Identification of Inadequate probe fits

Wideband reflectance measurements in young babies are sensitive to the quality of the acoustic seal, occlusion of the probe tip due to ear canal debris or contact with the ear-canal wall, and fussiness of the baby (AlMakadma & Prieve, 2021; Hunter et al., 2010; Merchant et al., 2010; Vander Werff et al., 2007). It is difficult to visually determine adequacy of probe fit due to the tiny diameter and length of the newborn ear canal, especially for screeners without audiologic training. Fortunately, WAI tests include complex acoustic responses measured at the probe tip (Keefe et al., 1993) such as absorbance, impedance magnitude, and phase that can be compared to the calibrated sound source to ascertain whether a technically adequate probe fit with sufficient energy transmission is present in the ear canal response (Voss et al., 2016). A blocked probe tip, either against the canal wall or due to material blocking the microphone port leads to a reduced absorbance close to zero. Leak effects in adult ears show an increase in absorbance between 0.1 and 0.3 kHz as the diameter of the leak increases (Groon et al., 2015), while in infants the primary frequency range of increased absorbance is 0.25 to 1 kHz (AlMakadma & Prieve, 2021).

Recommendations to identify inadequate probe fits, whether due to a leak or blocked probe tip, have been based on several different ambient absorbance wideband variables. These have included a negative equivalent volume of the ear, e.g., a volume less than −1.15 cm^3^ between 0.25 and 1 kHz (Keefe et al., 2000), negative impedance phase, e.g., phase below −0.11 cycles between 0.5 and 1 kHz (AlMakadma & Prieve, 2021), increased absorbance, e.g., reflectance less than 0.3 in low frequencies (Hunter et al., 2008) or alternatively absorbance greater than 0.7 at frequencies between 0.25 and 0.8 kHz (Aithal et al., 2015), or absorbance greater than 0.58 between 0.25 to 1 kHz (AlMakadma & Prieve, 2021). Previous studies of probe fit using wideband tympanometry have not been reported.

### 1.5. Study Objectives

The overall goal of this study was to assess wideband tympanometric (WBT) tests and wideband acoustic reflex thresholds (ART) in well-baby and NICU cohorts with three specific objectives: 1) Assess predictive accuracy for WBT and ART for conductive dysfunction in ears referring on the first or second stages of newborn hearing screening; 2) Identify inadequate tests likely due to probe blockages or leaks; and 3) Assess prediction models separately for well-baby and NICU screening outcomes. Methodological innovations in the present study were designed to address the study objectives, rigor, and reproducibility of the results.

## 2. METHODS

### 2.1 Participants

The protocol used in this study was approved by the Institutional Review Board of the participating hospitals. Informed consent was obtained from the parent(s) of all infants prior to any study procedures. Infants were eligible for enrollment after they received the first screening test in either the well-baby or NICU nurseries. The NHS protocol for the well-baby nursery was a two-stage procedure in which a TEOAE test (clicks at 80 dB SPL, Natus Medical, Inc.) was first completed in both ears 12-24 hours after birth. If the infant did not pass TEOAE in either ear, an automated ABR (clicks at 35 dB nHL, Natus Medical, Inc.) was completed as the second stage before hospital discharge. Recruitment was prioritized for infants who did not pass stage 1 screening to increase the failed screening sample for evaluation of sensitivity and specificity. The protocol for the NICU nursery was TEOAE followed by automated ABR, once the infant was medically stable and breathing room air, due to JCIH recommendations to include automated ABR to identify neural hearing loss. The WBT and ART test battery was performed as soon as possible after Stage 1 screening by a research assistant trained and supervised by an audiologist, usually the same day (85% of cases) but sometimes the following day due to the need to explain the study and allow parents time to decide whether to participate. Infants in the NICU were assessed by an audiologist.

After the initial TEOAE test was completed, and a parent consented to enroll in the study, infants were tested in both ears, as described in the Test Battery section. In the case of infants in the WBN, the infant was tested in the mother’s room or a procedure room adjacent to the nursery. In the case of infants in the NICU, testing was done in the baby’s isolette by an audiologist. The tester swaddled the baby and provided a pacifier, waited until they were quiet and not actively sucking, manipulated the outer ear to open it up, and selected a probe tip to fit as snugly as possible in the outer ear canal. The test battery commenced once an adequate seal was obtained. The pressure sweep was saved if judged of good quality, or discarded and repeated if the baby moved or made noise. This process continued at each step of the test battery, and pausing was available during the acoustic reflex testing if the baby began moving, yawning, sucking, or vocalizing.

Infants were classified as Pass or Refer for the TEOAE Stage 1 exam and the AABR Stage 2 exam. Demographics and risk factors for hearing loss are detailed in Table 1. The sex distribution was well-balanced between males and females. The racial makeup of the group was reflective of the greater Metropolitan area, with 58% White/Caucasian, 30% Black/African American, and 10% Other/Mixed, with few Asian or Hispanic/Latino participants. Due to the enrollment strategy to maximize screening referrals, there were more infants with risk factors than is typical, with about three times as many premature infants (32% of total) and NICU stays > 5 days (19%) than expected from the overall newborn population in the same hospitals. The TEOAE NHS failure rate (based on ears) was also higher than typical due to the enrollment strategy: 20% in the NICU sample and 47% in the well-baby sample. The 2-stage NHS failure rate was 10% in the NICU sample and 13% in the well-baby sample.

**Table 1.**
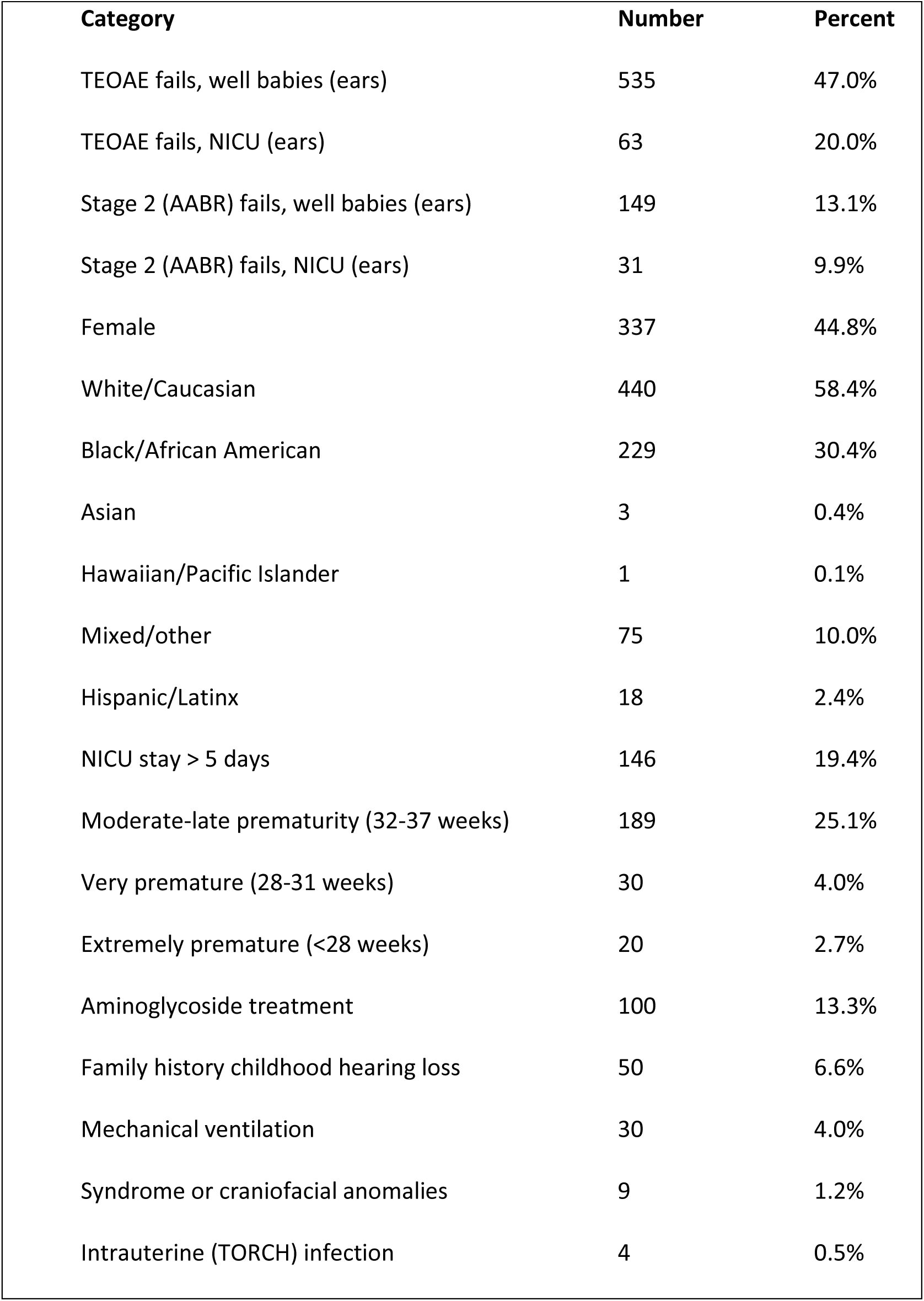
NHS Refer rates, demographics, gestational age at birth, and risk factors for congenital hearing loss. Total number of participants is 753.

### 2.2. WAI Test Battery

WB immittance data were acquired using a manufacturer prototype of an ear-canal probe used in the Titan device (Interacoustics, Denmark) and a AT-235 tympanometer (Interacoustics, Denmark) that was modified to allow custom software control of its pump and controller. The research methods followed those described by Keefe et al. (2015). All WAI tests were performed over a frequency range from 0.25 to 8 kHz. The acoustic response to a train of clicks with an inter-click interval of 46 ms was measured during the pressure sweep in each direction. The typical total SPL of the click train used to measure the WAI responses was 69 dB in both ambient and tympanometric tests. Silicone disposable probe tips were fit to the probe to provide a hermetic seal for pressurized measurements. The WAI test battery was composed of a downswept tympanometric WB test, followed by a WB ambient test, then an upswept tympanometric test, and lastly a WB acoustic reflex test. The pressure range was +200 daPa to −300 daPa in the downswept tympanogram and was −300 daPa to +200 daPa in the upswept tympanogram. The air pressure in the ear canal was monitored throughout testing, and the pump was automatically reset by the software to 0 daPa between each test. Preliminary testing in infants showed that starting the test battery with down-swept tympanometry confirmed whether the probe insertion was leak-free at the most positive air pressure of +200 daPa and minimized effects of ear-canal collapse on the subsequent ambient test. For this reason, the test battery order was kept constant. For the purpose of this report, the downswept tympanometric test, ambient test, and ART were used to develop predictive models and assess test performance. Preliminary analysis showed that the upswept test frequently resulted in ear canal collapse, and did not perform well for prediction, so it was excluded from further analysis.

### 2.3. WB Acoustic Reflex Test

The WB ART was completed last in the test series using the same probe and eartip used for other WAI tests, while maintaining its position in the ear canal unless refitting was necessary. Test procedures were fully described in Keefe et al. (2017) and Hunter et al. (2017). Briefly, ART test stimuli were sets of five clicks in a pulsed-activator stimulus set, where one click was delivered by the first receiver and four pulsed BBN activators were delivered by the second receiver. The initial, or “baseline,” click in each pulse-activator set relative to the post-activator clicks provided a set of four click-difference waveforms. The presence of a shift in the absorbed power level calculated from these click-difference waveforms relative to a criterion shift level specified in the above references defined a valid reflex, objectively determined by the automated test without input from the examiner. The stimulus set was ten activator levels in 5 dB increments ascending in level. Each activator level was specified by the SPL measured in a reference to a 2 cm^3^ coupler. ART responses were measured at the average of the TPP for the down-swept and up-swept tympanograms. The air pressure in the ear canal was continuously monitored during the ART test at TPP using a pressure sensor within the probe assembly. Whenever the measured air pressure deviated by more than 10 daPa from the desired TPP, the system paused the reflex test allowing the operator to adjust the probe fit to allow the pump to produce the desired pressure, and the test was re-started at the current activator level.

### 2.4. Calibration methods

Calibration was performed daily prior to testing using ambient pressure measurements in a pair of cylindrical tubes having lengths of 236.5 cm and 5.9 cm, and area of 0.178 cm^2^ (diameter 0.476 cm). Each tube was closed at its far end. The calibration procedure calculated the source pressure reflectance of the probe and the incident pressure spectrum of the click for frequencies between 0.2 and 8 kHz using the acoustic measurements in the tubes and by fitting the tube data to a model of viscothermal sound transmission in each tube. The tube area was used to calculate admittance in terms of reflectance. Once this calibration was completed, the electrical stimulus level of the click was maintained constant for all ear tests. Further detailed methods used to calibrate and measure WB tympanograms are described in Keefe et al. (2015).

### 2.5. WAI Analysis

Data were recorded using custom software running on a personal computer with a two-channel CardDeluxe sound card (22.05 kHz sample rate, 24-bit converters) and RS-232 serial port using custom software. All processing and subsequent analyses of WAI test data described below were performed using MATLAB (version R2022a) with its Statistics and Machine Learning Toolbox. Responses were analyzed in terms of absorbance and group delay (i.e., the negative phase gradient measured in units of time across frequency). WB test variables calculated from the WB sound pressure measurement at the probe tip included absorbance (A), group delay (D), complex admittance [any of magnitude (YM), phase (YP), conductance or real part (YR), susceptance or imaginary part (YI), sound pressure level (SPL), and absorbed power level (WL)]. The unit of each of the admittance variables YM, YR and YI was the acoustic mmho (CGS), and the unit of YP was degrees. The unit for absorbed (root mean-squared) power level (WL) was defined as WL=0 dB for a sinusoidal tone with SPL=0 dB acting on a conductance of 1 mmho (CGS). Responses were averaged into eleven half-octave frequency bins to reduce the data to a manageable number of points to be analyzed between 0.25 and 8 kHz.

### 2.6. WB variable names

Each WB variable was identified by one of: A, D, YM, YP, YR, YI, SPL or WL. Ambient WB variables were named by appending a lower-case ‘a’ to the WB variable name. For example, Aa represented a WB ambient absorbance response. Tympanometric WB variables from the down-swept WB tympanogram were named according to one of three times during the sweep: the initial positive-tail pressure (‘pt’), the TPP (‘tpp’), or the final negative-tail pressure (‘nt’). For example, YIpt denoted the WB tympanometric susceptance at the pt pressure. Data from the up-swept tympanogram was not used in the analysis because the canal often collapsed for the initial most negative air pressure and tended to remain collapsed until positive air pressure pushed it open at the end of the upswept test.

### 2.7. Probe fit exclusion criteria

Based on acoustic principles and studies of ear-canal acoustics reviewed in the Introduction section, low-frequency patterns of equivalent volume, admittance phase, and susceptance share common features. Susceptance at low frequencies (up to 1 or 2 kHz) is proportional to the product of equivalent volume and frequency and is also proportional to admittance phase. Excessively negative values of any of these variables indicate a mass-controlled admittance associated with the presence of an air leak. A complicating factor in neonatal ears is that susceptance can be slightly negative due to a finite mobility of the ear-canal wall at low frequencies (Keefe et al., 1993; Keefe et al., 2015). Thus, the exclusion criterion for susceptance should be based on excessively negative values and not simply a negative value close to zero. In contrast, in a valid test, a leak and blockage-free insertion of the probe tip into a neonatal ear canal (in the absence of wall mobility effects) would lead to positive values of equivalent volume, admittance phase, and susceptance at frequencies up to 2 kHz or more.

An adequate fit test was designed to identify both leaky and blocked types of probe fits. Based on preliminary analyses in a subset of WBN ears and prior research as reviewed in the introduction (Section 1.4), a criterion to classify a test as having an inadequate probe fit was based on an extremely negative susceptance or excessively low SPL suggesting an air leak and an excessively low absorbance (less than 0.005) suggesting a blocked probe. Because the absorbance level is 10 times the common logarithm of absorbance, the criterion of 0.005 is a 26 dB reduction from the maximum absorbance level of 0 dB. This test of probe fit adequacy was applied at frequencies between 0.5 and 2.8 kHz in the WB response, which is a sensitive bandwidth for both probe fit issues and evaluating middle-ear status (AlMakadma & Prieve, 2021). The impedance phase recommended in AlMakadma & Prieve was close to the susceptance selected for the criteria in the present study, and the 0.5-2.8 kHz bandwidth includes the bandwidth they recommended. The effect of excluding tests with inadequate probe fits based on these criteria was examined.

### 2.8. Newborn Hearing Screening exam groups

As described above, NHS exams were based either on the pass/refer results from a TEOAE only (Stage 1) or TEOAE followed by automated ABR (Stage 2). In the case of babies cared for in the NICU, they received both tests so they were assigned pass or refer groups independently, based on the TEOAE and automated ABR test results. The total number of participants was 753, and the total number of test ears was 1,452. Participants were randomly assigned to either model development (Group 1) or validation (Group 2) according to a 60:40 split. The development group was larger to facilitate the multivariate model design described below. The randomization was structured so that a close approximation to a 60:40 split was present for each sex and in each of the pass and refer sub-groups based on the TEOAE exam. Both ears of an infant were assigned to the same group, to avoid bias in the validation group. The relative numbers of ears in each group for each NHS exam are shown in Table 2, which also shows the number of ears in the Pass sub-group (NN) and the Refer sub-group (NI), and the numbers identified as outliers based on probe-fit criteria. Further details are provided in the Results (section 3.1.3).

**Table 2.**
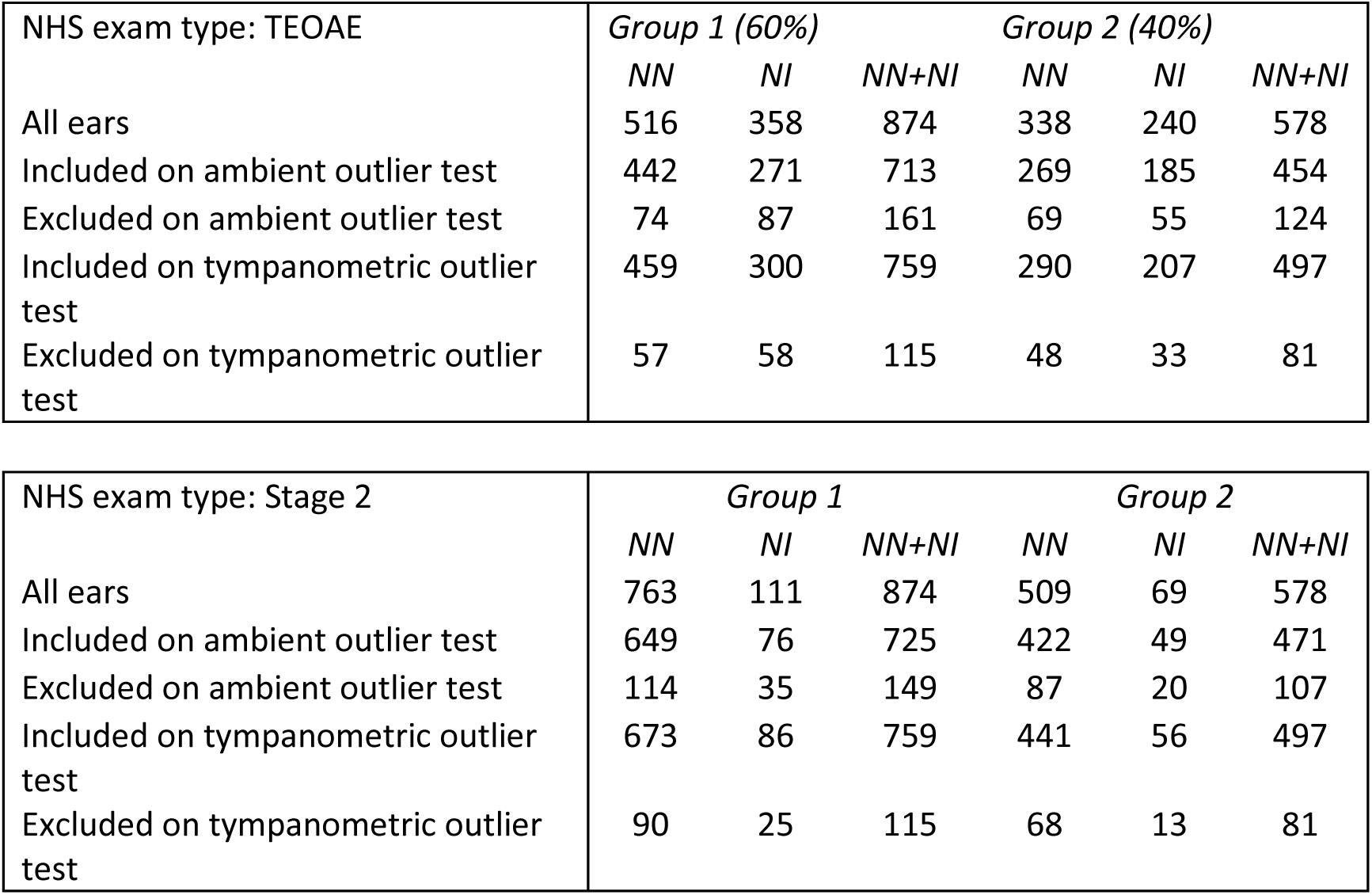
Number of test ears with NN as number of pass ears and NI as number of refer ears (or number impaired) on the respective NHS exams TEOAE and Stage 2 in Group 1 and Group 2. The number of outliers included or excluded are shown for the WB ambient and tympanometric tests.

### 2.9. Overview of statistical model analyses

The analysis procedures were devised according to procedures used in previous studies described in the Introduction with novel procedures added. Separate predictive models of NHS exam outcomes were constructed based on: (1) NHS exam type (TEOAE, Stage 2), (2) WAI test type (ambient, tympanometric), (3) whether data from an ART test at TPP were included in the model, (4) whether inadequate tests were excluded from the model inputs, and (5) whether the model inputs were a selected set of WAI variables or principal components (PCs) calculated from a PCA of the set of WAI variables. Further methodological details are specified in the Results (section 3.1.4) because the results from one step of the analysis were used to design and conduct the next step of the analysis.

Exploratory data analyses consisting of the mean, standard error (SE) of the mean, and Tukey-style box-and-whisker plots were completed on all subject data and separately for data in Groups 1 and 2. Separate exploratory analyses were completed based on the above alternatives in steps (1) and (2), and for the ART data used in step (3). Group data were similar in all ears compared to the subset of Group 1, which lent confidence on using Group 1 as the development group on which to construct the predictive models. Outlier tests for step (4) were devised based on box-and-whisker analyses of SPL and susceptance and on values near zero of the absorbance.

The ability of all WAI variables (including ART) to classify ears that passed or failed the NHS exam was quantified in terms of each of their univariate AUCs. A subset of these WAI variables with the largest univariate AUCs were selected as potential inputs to multivariate predictors to classify NHS outcomes. A PCA was calculated for these WAI variables in Group 1 to provide a set of uncorrelated principal component (PC) inputs for an alternative multivariate predictor in step (5) above. Small logistic-regression models were initially constructed to investigate which pairs or triplets of WAI variables led to model outputs that were most accurate based on AUC. Information from these preliminary analyses informed the design of the final multivariate models and the choices of input WAI variables. The number of variables included was limited to reduce the possibility of overfitting the model. Each final model was a stepwise general linear model (GLM) with a binomial distribution to best classify the NHS exam outcome of each ear using a logistic regression of the model outputs. Burnham and Anderson (2004) recommend using either the Akaike Information Criterion (AIC) or Bayesian Information Criterion (BIC), whichever is most predictive, to improve the accuracy of a model, noting that smaller models tend to generalize to new groups better than overly large models. The ability of each model to classify ears was quantified by its AUC value on Group 1. The output of the development Group 1 analyses was a set of multivariate models having varying number of WAI inputs depending on the criterion AUC. The generalizability of each of these models was assessed by using data from validation Group 2 as the model inputs. The multivariate model having the maximum AUC on the Group 2 dataset was defined as the best model. Evidence of model overfitting was identified if the AUC for Group 1 was much larger than the AUC for Group 2.

## 3. RESULTS

### 3.1 Ambient Test Data

#### 3.1.1 Mean and SE

The results for Group 1 are shown in Fig. 1 (mean ± one SE values of absorbance, group delay, admittance, SPL, and WL) for TEOAE pass and refer sub-groups. There were pronounced mean differences in pass and refer sub-groups with the largest effect sizes over mid-range frequencies for absorbance and conductance (Fig. 1A). Effect size was measured as the ratio of the mean difference between the sub-groups and the pooled SE (Cohen, 1988). A distinctive feature of the mean difference of group delay (Fig. 1B) was that it was largest at lower frequencies (0.5-0.7 kHz) compared to mid-frequencies in other responses, except for susceptance. However, SEs were larger relative to the mean difference than those observed for other variables, i.e., its effect sizes were smaller. The mean difference between pass and refer ears for absorbance and conductance (Fig. 1A and C) was largest around 1.4-2 kHz. Susceptance (Fig. 1D) was higher for Pass ears from 0.71 to 1 kHz, while it was higher for Refer ears at frequencies above 1.4 kHz. The mean difference for power level (Fig. 1F) was largest at 2 kHz and was negligible at and below 1 kHz. Although not typically included as a WAI variable, the SPL (Fig. 1E) was larger in the referred sub-group at lower and mid-frequencies.

**Figure 1.**
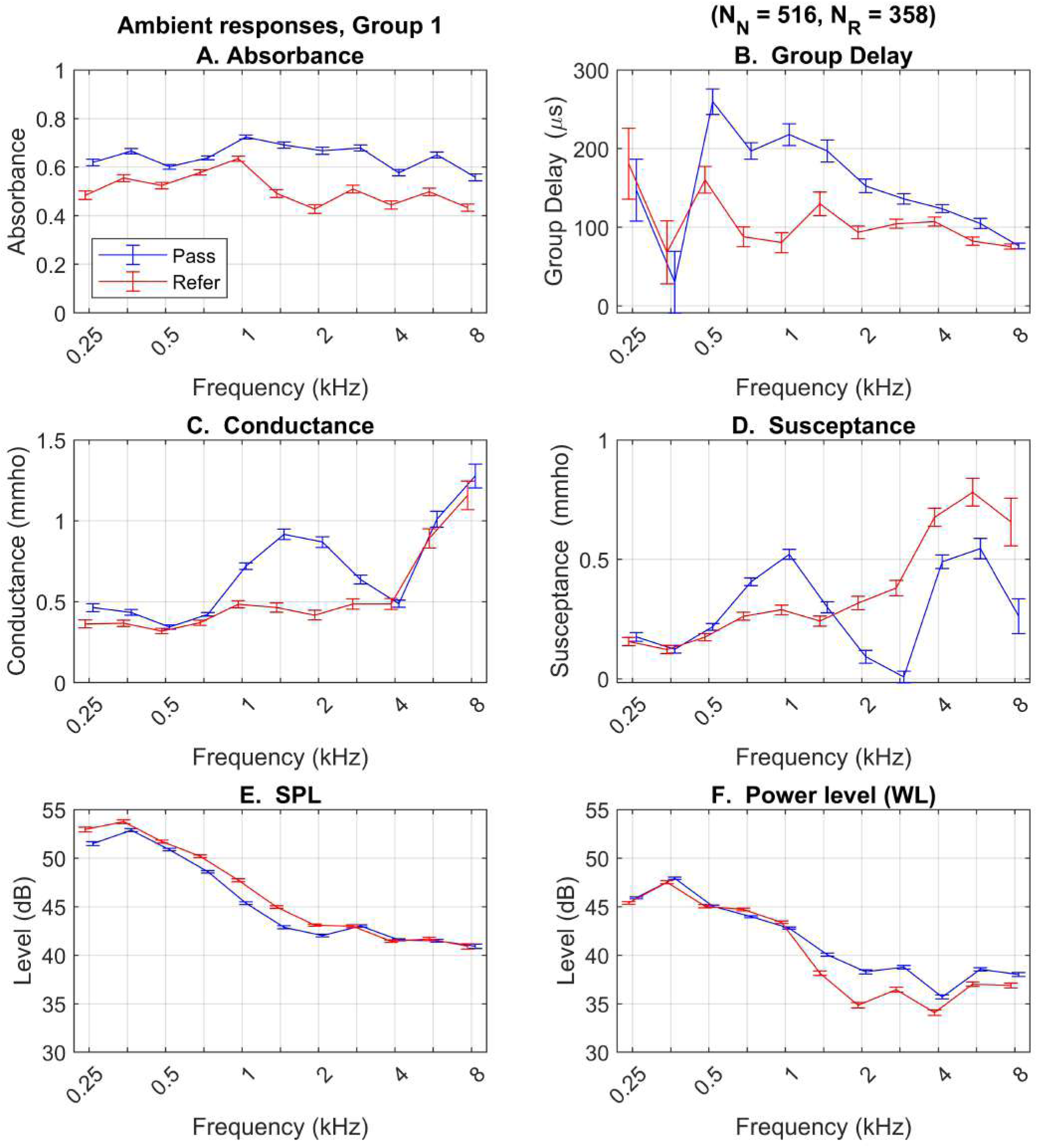
Mean + 1 SE are shown for each ambient WB variable in Group 1 for pass and refer groups defined by the TEOAE exam. Conductance shown as Real Admittance and Susceptance as Imaginary Admittance.

#### 3.1.2 Univariate AUCs

Univariate (non-parametric) ROC analyses were performed to classify ears as pass or refer on the TEOAE exam across ambient WAI variables in Group 1 at each frequency bin, and are shown in Fig. 2. Univariate AUCs ranged between 0.5 (chance performance) up to 0.71. The frequencies at which a particular WAI variable had the largest AUC values were considered as potential candidates in multivariate predictors. Overall, the largest AUCs occurred at frequencies between 0.7 and 2.8 kHz, which were consistent with the largest group differences in the mean results in Fig. 1. The most accurate predictors of TEOAE status for ambient tests occurred at these frequencies: Absorbance at 1.4-2.8 kHz, power level (WLa) at 2 and 2.8 kHz, SPL at 1 kHz in the top panel, admittance magnitude (YMa) at 1-1.4 kHz, admittance phase (YPa) at 2-2.8 kHz, conductance (YRa) at 1-2 kHz, and susceptance (YIa) at 0.7-1 kHz and 2.8 kHz in the bottom panel. Qualitative patterns for the Stage 2 exam were similar to those for the TEOAE exam. Because the AUCs for group delay were smaller than for other WAI variables, due to higher variability, group delay was omitted from further model analyses.

**Figure 2.**
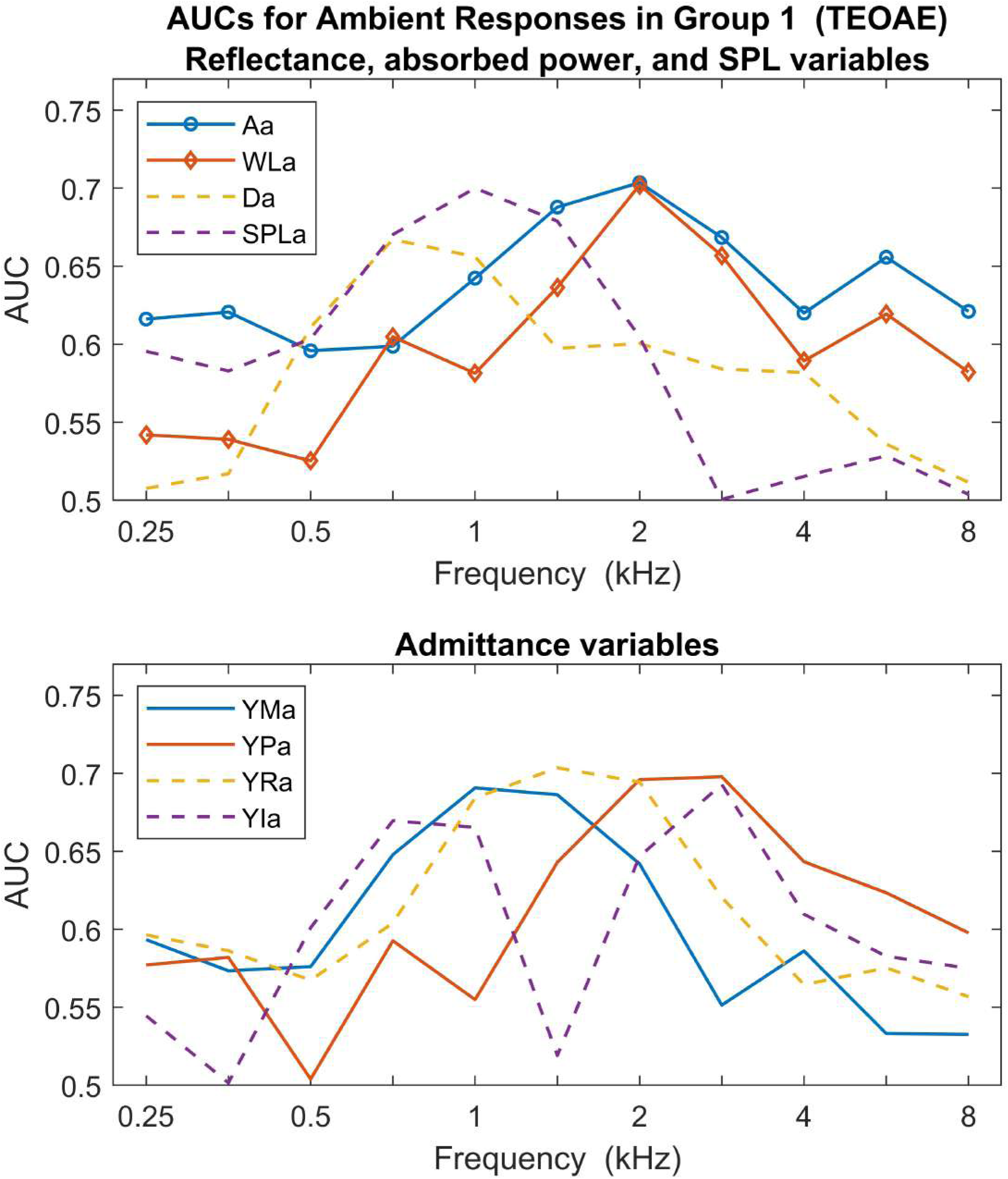
Univariate AUCs are shown for all ambient responses in Group 1, with variable type defined in the legends of the top and bottom panels. Top legend variables: absorbance (Aa), absorbed power level (WLa), group delay (Da), sound pressure level (SPLa). Bottom legend variables: admittance magnitude (YMa), admittance phase (YPa), conductance (YRa), susceptance (YIa).

The WB ART univariate AUC for the TEOAE (i.e., Stage 1) exam was 0.702 in Group 1 and 0.702 in Group 2. Its univariate AUC for the Stage 2 exam was 0.707 in Group 1 and 0.723 in Group 2. Thus, the ART response had predictive accuracy similar to the best of the frequency-specific ambient WAI responses.

#### 3.1.3 Ambient Exclusion Classifier

As described earlier, the WB variables selected to identify inadequate tests were SPL, susceptance (YIa) and absorbance. A common method to exclude extreme outliers is the lower whisker value of a distribution of test responses that is the greater of the minimum value across all responses and 1.5 times the inter-quartile range (IQR) below the 25th percentile. This lower whisker value was used as the exclusion criterion value in the present study. A test was considered inadequate in a frequency bin if its SPL or YIa was less than the exclusion criterion value at each frequency. Such extreme results were theoretically due to a leaky probe fit. A test was also considered inadequate in a frequency bin if its absorbance was less than 0.005, which corresponds to an absorbance level reduced by 26 dB relative to an absorbance of 1 (at 0 dB). Such results were likely due to a blocked probe tip either with debris or against the canal wall. Either type of result might also occur due to errors in calibrating the system for WB admittance and reflectance measurements, hardware issues, or testing difficulties related to the behavioral status of the infant. The numbers of ambient WB responses that were included and excluded in subsequent multivariate model analyses are listed in Table 2 for the NHS types TEOAE and Stage 2.

The criterion values for the ambient exclusion criteria are listed in Table 3 for frequencies between 0.5-2.8 kHz. In summary, an ear test was initially classified as inadequate based on any of these three variables at any frequency. To achieve a balance between the need to construct the model with a large sample population against the need to exclude inadequate measurements, the number of frequency bins used to identify inadequate tests was restricted. Preliminary analyses showed that the univariate AUC of any single WAI variable was small at low frequencies of 0.25 and 0.35 kHz and at high frequencies of 8, 5.7 and 4 kHz, so these bins were not included in the outlier test. The resulting final form of the outlier test considered the most relevant bin frequencies for prediction value (0.5, 0.7, 1, 1.4, 2, and 2.8 kHz, see Table 3). The relative proportion of outliers was larger for refer ears than for pass ears in each group for each NHS type. Outliers were more often classified based on absorbance close to zero than on the other two leak-detection variables, especially above 1 kHz, indicating a likelihood of a blocked probe tip.

**Table 3.**
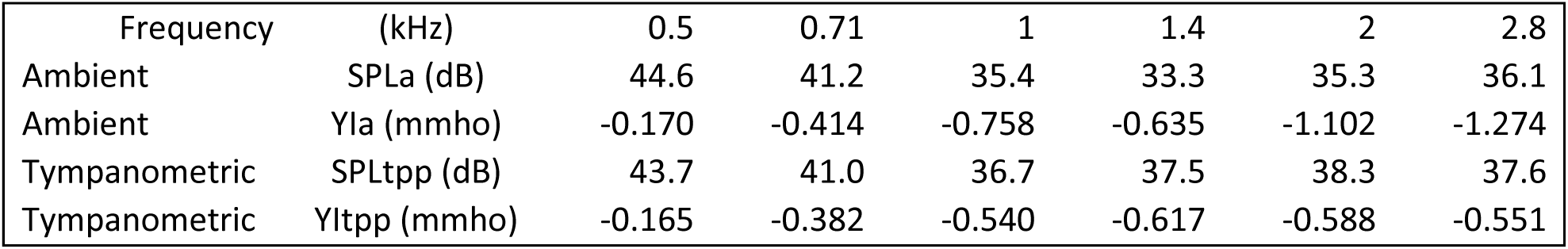
Criterion values for outlier tests based on lower whisker values in boxplots of ambient SPL (SPLa), ambient susceptance (YIa), tympanometric SPL at TPP (SPLtpp) and tympanometric susceptance at TPP (YItpp). Criterion values are defined at frequencies from 0.5 to 2.8 kHz. Any test value below any of these criteria as an outlier on the ambient and tympanometric tests, respectively.

#### 3.1.4 Multivariate Model Development

A stepwise general linear model (GLM) with a binomial distribution was used on Group 1 tests in both the ambient and tympanometric test analyses, which resulted in a multivariate logistic regression to calculate the probability that each ear test in Group 1 was a pass or refer on an NHS exam. The initial input variable set included a constant intercept term, a linear term for each predictor, and all products of pairs of distinct predictors to account for interactions. GLM analyses were performed in Group 1 for all ear tests and those for which outliers were excluded.

The variables included in a select multivariate model were only those whose AUC in Group 1, termed AUC1, exceeded a minimum value (AUCmin). Models with progressively more ambient test inputs were calculated by varying AUCmin from 0.70 down to 0.62 in steps of 0.01. The upper limit was selected based on the few univariate AUCs that exceeded 0.70 for NHS types TEOAE (in Fig. 2) and Stage 2, and the lower limit was adjusted to limit the total number of input predictors in any model.

All models were analyzed using either AIC or BIC as the model criterion to limit the number of input predictors in the final model to avoid over-fitting the data. The stepwise GLM model procedure for Group 1 halted when the removal or addition of any input variable did not decrease the value of AIC or BIC. Any coefficient with p>0.05 was removed from the model to improve and simplify its structure. The final pruned model was selected based on the minimum value across the simplified models based on the consistent AIC value. The AUC2 was calculated in the Group 2 evaluation phase in terms of the GLM determined on Group 1 data. This AUC2 was the final measure of test performance. The predictive results for the best multivariate model are shown in the top half of Table 4 in terms of the final measure AUC2 on the data from Group 2. Each row corresponds to a choice of NHS exam, whether all tests were included or only those ears left after excluding outliers, whether the ART was included in the GLM, the choice of model criterion, and the value of AUC1 calculated for the data in Group 1. The right-most column shows the logit function of the best GLM in Wilkinson notation (Wilkinson and Rogers, 1973). It is evident from the logit function whether the best set of test variables is used with Aa and WLa or the two largest PCs named PCa and PCb. Detailed model specifications are available in Supplement Appendix A.

**Table 4.**
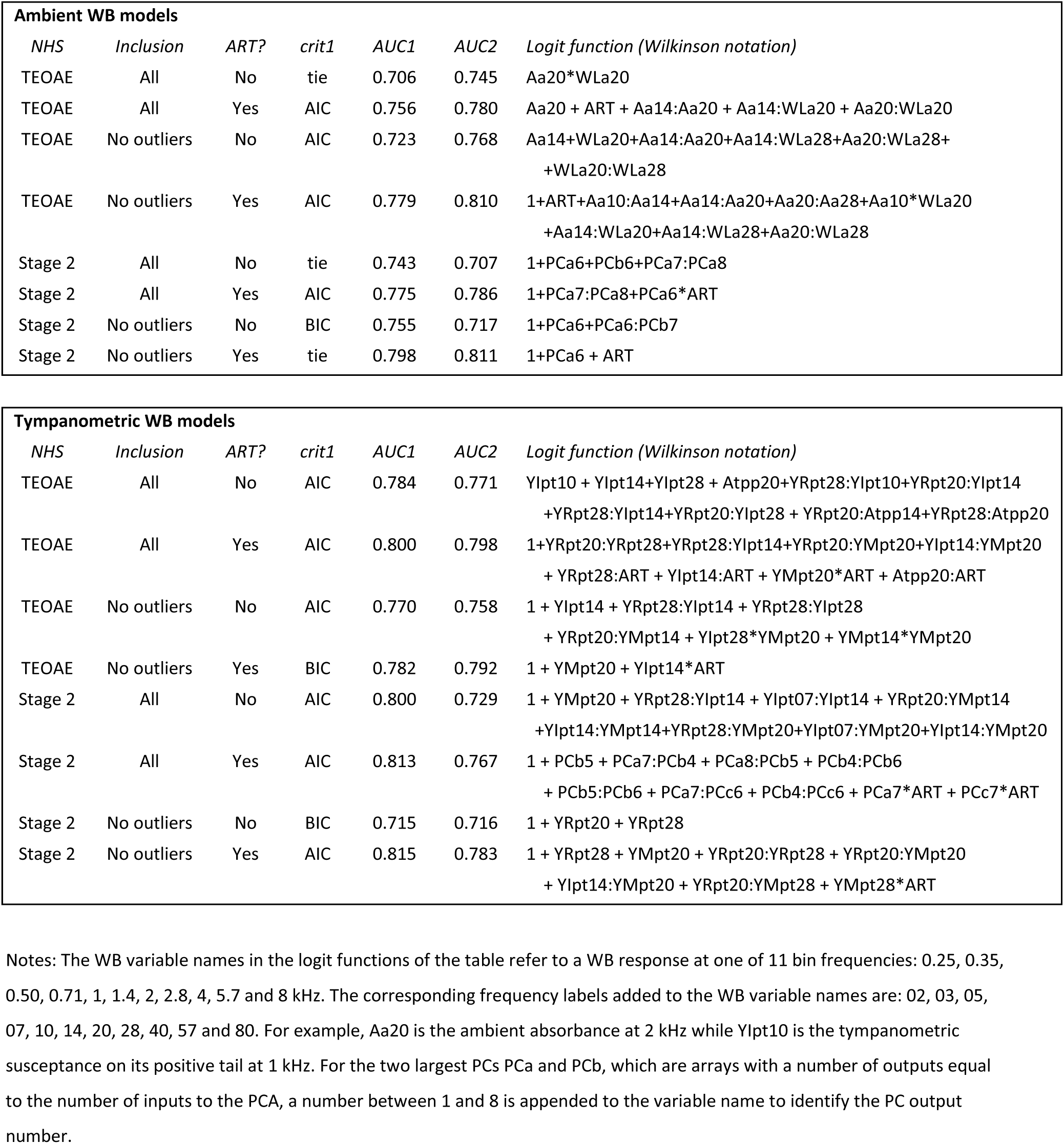
Results for Ambient (top) and Tympanometric (bottom) WB models. Columns include NHS exam type, whether all ears were included or those ears with outliers excluded, whether ART test was in model, model criterion for best model (crit1), best AUC for evaluation group (AUC2) along with the AUC calculated in its model development group (AUC1), and logit function of best model.

Considering performance on the TEOAE exam (Table 4), AUC2 was slightly larger than AUC1 on all four test conditions, which showed better than expected generalization of the predictors calculated for Group 1 on the data of Group 2. The models that included ART consistently performed better than models without ART, i.e., compare AUC2 in row 2 versus row 1 and in row 4 versus row 3. This shows the advantage of predicting TEOAE status with a combination of a WB frequency-specific test and the ART. The models that excluded outliers performed slightly better than those with all data, i.e., compare AUC2 for row 3 versus row 1 and row 4 versus row 2. This shows that the criteria to detect outliers improved test performance. The number of terms in the best logit functions were larger for models that excluded outliers (rows 3 and 4) than those that analyzed all variables (rows 1 and 2). The logit functions across all four TEOAE conditions contained Aa and WLa variables between 1 and 2.8 kHz. The best models used AIC in each of the four rows versus BIC in one row. Models with the original set of variables always performed better than those in which a PCA was performed.

Considering performance on the Stage 2 exam, the pattern of results was similar in most comparisons to those on the TEOAE exam. Namely, AUC2 was larger than AUC1 on two of the four test conditions, which again showed excellent generalization of the model to a novel set of test data. Models that included the ART performed better than those that did not. Models that excluded outliers performed slightly better than those with all data. The major difference between the Stage 2 and TEOAE results was that all the best Stage 2 models included a PCA, which was calculated from the underlying Aa and WLa variables within a bandwidth between 1.4 and 2.8 kHz (see Supplement Appendix A.2). The PC denoted PCa6 in each of these models explained the largest amount of the variance (84 and 92%) in the original PCA. The definition names of other PCs are listed in the Notes section of Table 4.

#### 3.1.5 WBN compared to NICU comparisons

An issue considered following these main analyses is that NICU infants have more immature middle ears at birth, and greater risk of both conductive and SNHL. This higher-risk NICU group has not previously been compared to WBN infants on WB test performance. Thus the subgroups of WBN and NICU infants in the evaluation sample (Group 2) for the best ambient and tympanometric multivariate models were separately analyzed and compared. The best multivariate predictor models were reassessed both with and without the inclusion of outliers, and with and without ART as before, and results are shown in Table 5. The ART univariate test is also shown in Table 5 with and without outliers. The ambient WB results for the TEOAE NHS exam showed larger AUC2 in WBN ears and smaller AUC2 in NICU ears. Differences were most apparent for the TEOAE test (in WBN group1 only), where the fail rate was higher than in the NICU group (fail rate shown in Table 1). For the Stage 2 exam, slightly larger AUC2 was also found in WBN ears and smaller AUC was found in NICU ears. For ambient WB models considering results on Stage 1 and 2 combined, 6 out of 8 comparisons were higher by at least 0.10 for AUCs in the WBN group compared to the NICU group, and the AUCs in 2 of 8 comparisons were nearly equal. There was a definite benefit to including the ART test in NICU ears. For the Stage 2 test, when ART was included, there were no differences in the WBN and NICU groups.

**Table 5.**
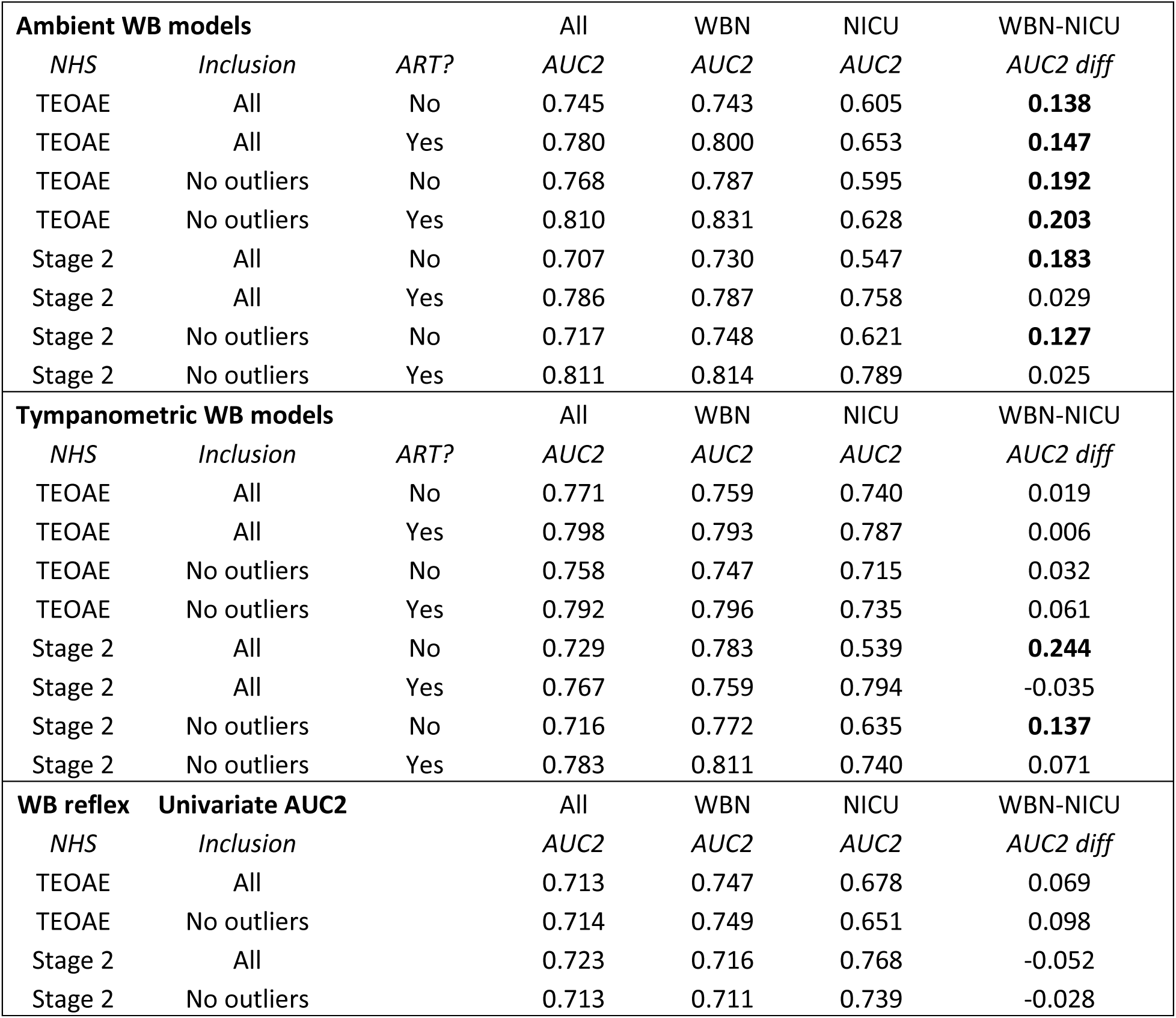
Results for Ambient (top), Tympanometric (center) WB models and WB Reflex univariate AUCs for all ears, and for the well-baby nursery (WBN) and NICU separately. Columns include NHS exam type, whether all ears were included or those ears with outliers excluded, whether ART test was in model, and AUCs for the evaluation group (AUC2). The last column shows the WBN-NICU difference in AUC and is bolded if the difference is greater than 0.1).

### 3.2 Tympanometric Test Data

#### 3.2.1 Mean and SE

The mean and + SE responses for the pass and refer ears on the TEOAE exam in Group 1 for tympanometric absorbance and group delay are shown in Fig. 3. Three air-pressure conditions are shown: positive-tail pressure (Ppt) at +200 daPa, TPP, and negative-tail pressure (Pnt) at - 300 daPa. The responses varied between pressure conditions. The mean absorbance at TPP (Fig. 3B) was larger in the pass ears with the largest mean differences between 1.4 and 2.8 kHz. A large mean difference in group delay at TPP (Fig. 3E) was apparent at 1 kHz. The mean absorbances at Pnt (Fig. 3C) had the smallest values compared to those at TPP and Ppt, and similarly for mean group delay at Pnt (Fig. 3F) except at 0.25 kHz. The mean absorbance at Ppt (Fig. 3A) increased with frequency from 0.25 to 0.7 kHz, with no difference between pass and refer ears between 0.25 and 1.4 kHz. The mean absorbance at Ppt in the pass ears was much larger than in refer ears at and above 2 kHz. The mean group delay at Ppt (Fig. 3B) was larger in pass ears at frequencies between 1.4 and 2.8 kHz, but the effect size was smaller than for absorbance due to the larger SEs in group delay.

**Figure 3.**
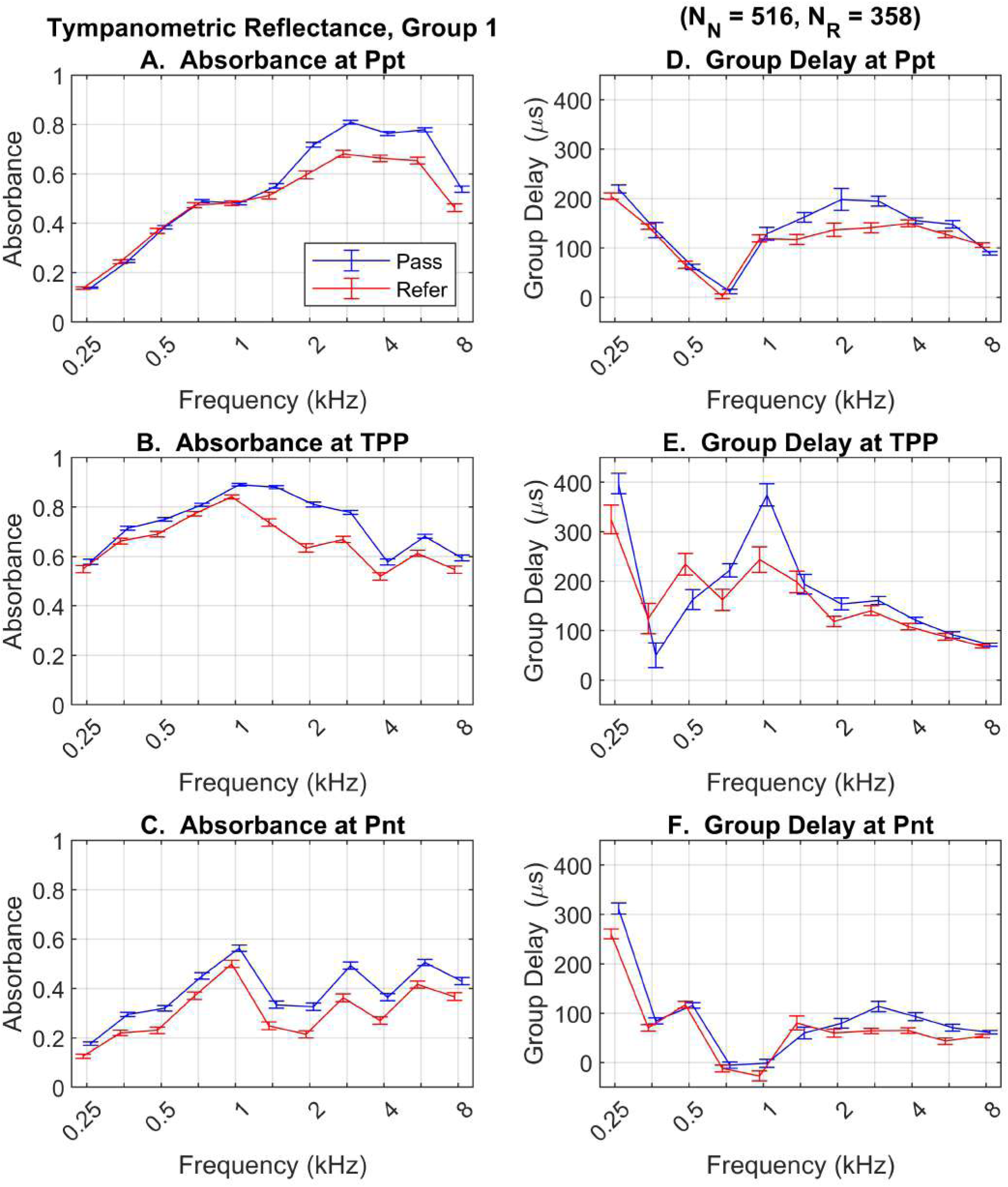
Mean + 1 SE are shown for tympanometric absorbance (left panels) and tympanometric group delay (right panels) in Group 1 for pass and refer groups defined by the TEOAE exam. Curves for Ppt (top row), TPP (middle row), and Pnt (bottom row).

The mean + 1 SE of the tympanometric admittance magnitude and phase are shown in Fig. 4. A clear result is the large SEs for YM in the pass ears at Pnt (Fig. 4C), coupled with the much smaller mean YM in the refer ears. This pattern is explained by the fact that negative air pressure (with respect to atmospheric room pressure) was associated with a collapse of the immature ear-canal walls in some ears, thus the large SE in the pass group may be explained by some ears with open canals and others with collapsed canals. The mean admittances at Ppt and TPP for YM (Figs. 4A-4B) and YP (Fig. 4D-4E) showed mean differences with large effect sizes at subsets of frequencies between 1.4 and 4 kHz, and no mean differences at lower frequencies. The mean YM was larger at Ppt than at TPP for all frequencies. The mean YP at TPP was non-negative at all frequencies, which showed a stiffness-controlled behavior.

**Figure 4.**
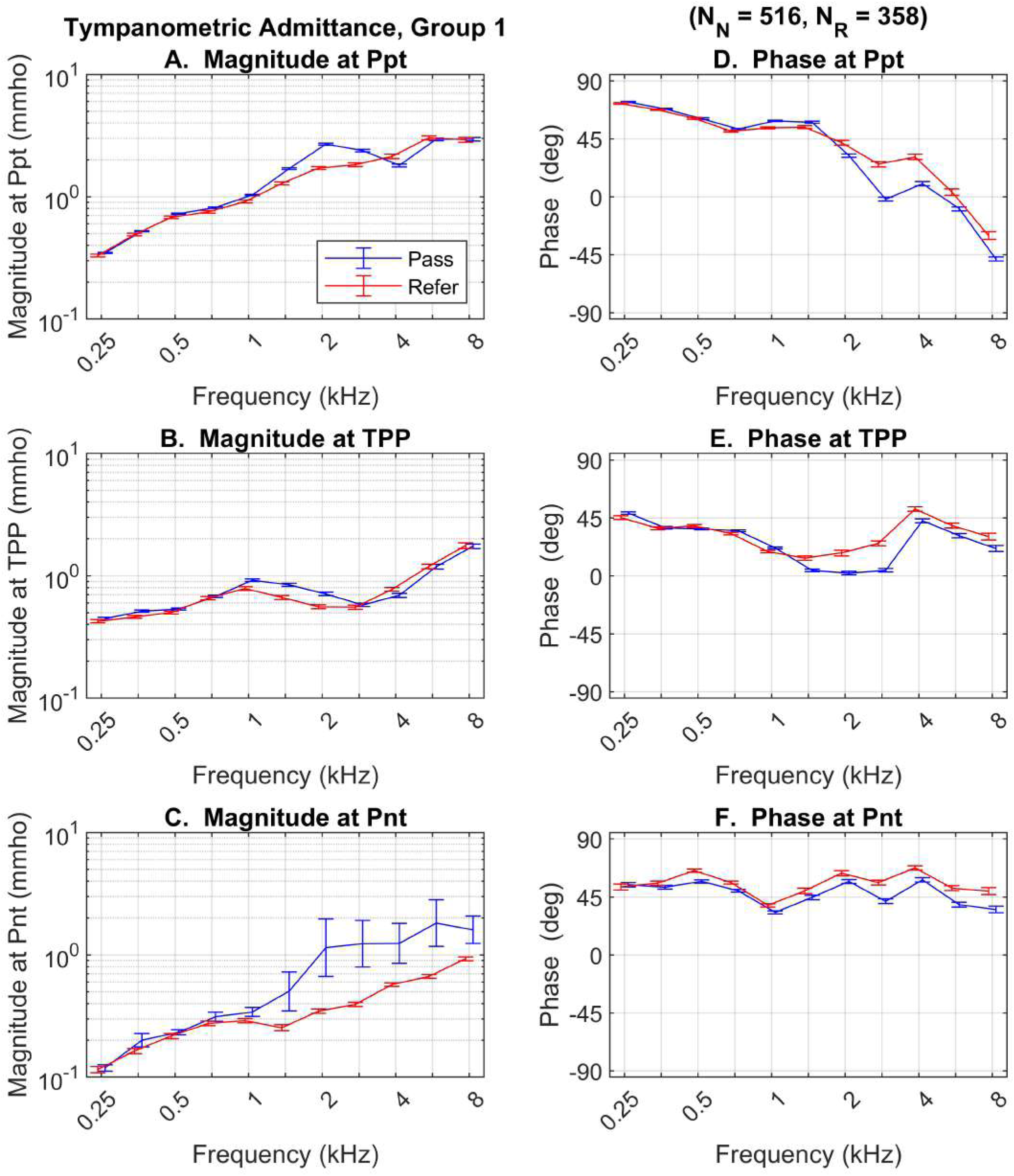
Mean + 1 SE are shown for tympanometric YM (left panels) and tympanometric YP (right panels) in Group 1 for pass and refer groups defined by the TEOAE exam. Curves for Ppt (top row), TPP (middle row), and Pnt (bottom row).

#### 3.2.2 Univariate AUCs

Univariate AUCs for tympanometric variables at selected air pressures (pt, TPP, nt) are shown in Fig. 5. AUCs were close to chance performance for variables at the nt pressure (not shown), so those variables were omitted from multivariate analyses. The largest AUC occurred for Atpp at 2 kHz, which was not obvious given that the corresponding mean difference in Atpp in Fig. 3A showed large effect sizes between 2 and 8 kHz. Because the univariate AUCs of WL and SPL were small, these variables were omitted from any multivariate predictors. The largest AUCs occurred for real and imaginary admittance variables for the positive-tail pressures (YRpt and YIpt, middle panel Fig. 5) at frequencies between 1.4 and 2.8 kHz. The largest AUCs occurred again for admittance magnitude and phase variables (YMpt and YPpt, bottom panel Fig. 5) at the positive-tail pressure at frequencies between 1 and 2 kHz.

**Figure 5.**
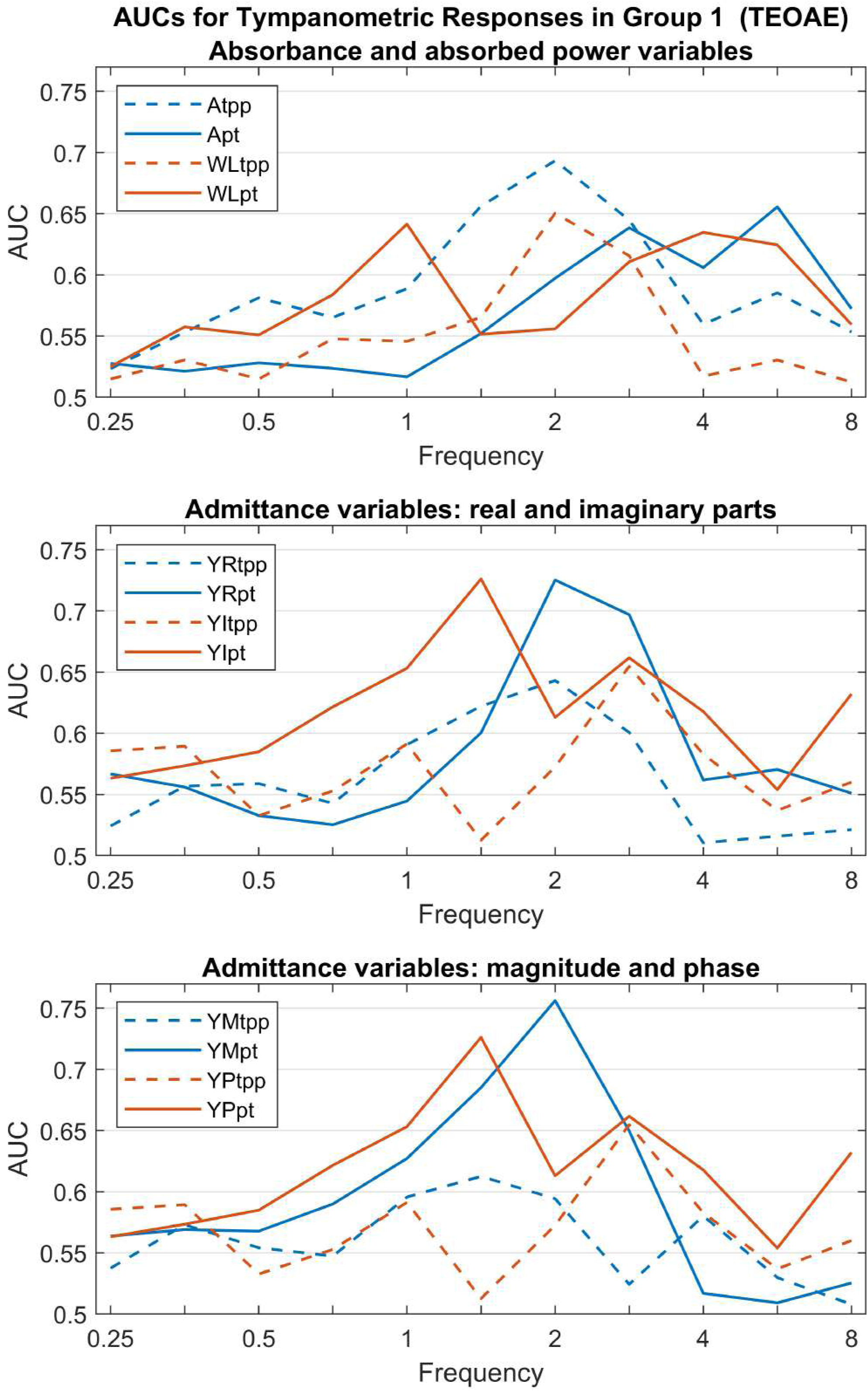
Univariate AUCs are shown for all tympanometric responses in Group 1, with variable type defined in the legends of each panel. Top legend variables: absorbance at TPP (Atpp), absorbance at Ppt (Apt), absorbed power level at TPP (WLtpp), absorbed power level at Ppt (WLpt). Middle legend variables: conductance at TPP (YRtpp), conductance at Ppt (YRpt), susceptance at TPP (YItpp), susceptance at Ppt (YIpt). Bottom legend variables: admittance magnitude at TPP (YMtpp), admittance magnitude at Ppt (YMpt), admittance phase at TPP (YPtpp), admittance phase at Ppt (YPpt).

#### 3.2.3 Tympanometric Exclusion Classifier

Different ambient and tympanometric classifications were needed because these were two separate tests. The test to identify tympanometric ear tests as inadequate was based on extremely low values of absorbance, SPL and susceptance at TPP. The criterion values of each response to classify a test as inadequate were defined in a manner analogous to those described above for ambient tests: absorbance at TPP less than 0.005, and values of SPL and susceptance at TPP below their criterion values defined in terms of their corresponding lower whisker values in Table 3.

Tympanometric variable classifications were performed at intermediate frequencies from 0.5 kHz to 2.8 kHz. The variable set at TPP was used to identify inadequate tests in an intermediate point in the downwards sweep. As for the ambient test, the numbers of tympanometric WB responses that were included and excluded in subsequent multivariate model analyses are listed in Table 2 for the NHS types TEOAE and Stage 2. Across both groups (independent of NHS exam type), 13.5% of ears were excluded. This was less than the 19.6% of ears excluded in the ambient test. The relative proportion of outliers was larger for refer than for pass ears for three of four cases in each group for each NHS type. Far fewer ear tests were excluded as outliers when absorbance, SPL, and susceptance at Ppt were used in the tympanometric outlier test rather than those at TPP. This was because the ability to pressurize to the maximum air pressure in the ear (200 daPa) was a strong indication that the probe fit was leak-free on the downswept test.

#### 3.2.4 Multivariate Model Development

Based on the univariate AUC analyses, the set of potential input variables to a tympanometric multivariate model were winnowed down to subsets of absorbance at TPP and admittance variables at Ppt. The final multivariate models were constructed across these subsets of WB tympanometric variables: (1) YRpt+YIpt+YMpt, (2) YRpt+YIpt+Atpp, and (3) YRpt+YIpt+YMpt+Atpp. All sets contained YRpt and YIpt, which was the best performing set of two variables. The multivariate models of tympanometric WB responses were constructed and analyzed on Group 1 data in an analogous manner to the ambient WB models, aside from changes specific to the larger univariate AUCs obtained for tympanometric variables. The range of AUCmin in tympanometric models was examined from 0.72 down to 0.64 in steps of 0.01, and the three subsets of tympanometric variables in the above paragraph were used rather than the one subset of ambient variables.

#### 3.2.5 Multivariate Models

The predictive results for the best multivariate model using the tympanometric WAI data are shown in Table 4 in terms of the final measure AUC2 on the data in Group 2. The corresponding detailed model specifications are available in Supplement Appendix B. Considering performance on the TEOAE exam, AUC2 was nearly identical to AUC1 on the four test conditions. These results demonstrate satisfactory generalization of the ability of the best GLMs to classify hearing status in a novel group. Models that included ART data performed better than models without. This again showed the advantage of predicting TEOAE status with a combination of a WB frequency-specific test and the ART. The models excluding outliers did not perform better than those with all data. The logit functions across all four TEOAE conditions contained tympanometric variables only between 1 and 2.8 kHz. Models with the original set of variables always performed better than those in which a PCA was performed. The best model in Table 4 that contained all four input tympanometric variables (YRpt, YIpt, YMpt, Atpp) was the model for all ears that also included the ART test with an AUC2 of 0.798.

Considering performance on the Stage 2 exam, the pattern of results was similar to those on the TEOAE exam. AUC2 was nearly always the same or slightly smaller than AUC1, which showed good generalization of the model to a novel set of test data despite the smaller number of refer ears on the Stage 2 exam relative to the TEOAE exam (see Table 2). Models that included the ART consistently performed better than those that did not. Models that excluded outliers performed about the same on AUC2 as those with all data.

#### 3.2.6 WBN compared to NICU comparisons

Performance on the best tympanometric WB models, and on the ART test alone compared for the WBN group and the NICU group and is shown in Table 5. Differences between the WBN and NICU groups were small, except for cases in which Stage 2 failures were the outcome, and the ART test was not included. In the NICU group, the best predictors consistently included ART and excluded outliers. AUC2 was similar in WBN and All ears. The largest difference was in Stage 2, whether outliers were included or not, and without the ART test (AUC2 of 0.729 for All ears and 0.783 in WBN). AUC2 was smaller in the NICU group than All ears for all models with no ART test. AUC2 exceeded 0.7 in NICU ears only when ART data were included. This is the same pattern as the results of the univariate AUC2 of the ART test, especially in the Stage 2 exam.

## 4 DISCUSSION

This study had a primary overall purpose to develop multivariate models of WAI to predict middle-ear dysfunction in newborn ears that referred on the first and second stages of hearing screening, considering criteria for inadequate probe fit. The main findings of this study concerned multiple WBT and ART variables that were evaluated for their ability to predict conductive dysfunction in both well and NICU newborns, with and without exclusion of tests likely to be affected by probe leaks or blockages.

### 4.1 How well did univariate ambient tests perform?

Pass versus refer differences in ambient absorbance across frequency were similar to previous studies (Aithal et al., 2015; Hunter et al., 2010; Myers et al., 2018; Sanford et al., 2009b) with the greatest differences and effect sizes falling in the mid-frequency range (1.4-2 kHz). Although these results showed overall higher absorbance and a flatter shape in both pass and refer groups than some previous studies, the group difference was similar to previous studies (Aithal et al., 2017; Aithal et al., 2013; Hunter et al., 2010; Sanford et al., 2009a). Other WAI variables used herein have been rarely studied in newborns. These new variables contributed extra information, improving the predictive models. Univariate AUCs were largest (around 0.7) for the TEOAE test in the ambient test for absorbance at 1.4-2 kHz, absorbed power level at 2 kHz, SPL at 1 kHz, conductance at 1.4 kHz, and admittance phase at 2-2.8 kHz. The univariate AUCs were similar to those reported by Aithal et al. (2015) for ambient absorbance compared to automated ABR or TEOAE NHS groups using an earlier version of the WB system employed in the current study. While group differences occurred in the same frequency region as previous studies, the univariate AUCs obtained in TEOAE and automated ABR groups were lower than those reported by three studies using concurrent DPOAE as a reference standard (Sanford et al., 2009; Hunter et al., 2010; Myers et al., 2018).

### 4.2 How well did univariate tympanometric tests perform?

Summarizing across all AUCs in Fig. 2: (1) the best univariate predictors occurred at frequencies between 1 and 2.8 kHz corresponding to a range of large mean differences in the pass and refer groups for many variables in Figs. 3-4, (2) the absorbance at TPP was the best predictor of any variable across absorbance, absorbed power level and SPL, and (3) the admittances at positive-tail pressure were better predictors of TEOAE status than those at TPP, while some of these were better univariate predictors than absorbance at TPP. These findings suggest that the functional status of the tympanic membrane plays a key role in the information captured in the WAI responses between 1 and 2.8 kHz. The advantage of using tympanometric variables at the positive pressure tail may be specific to newborns and young infants, as positive air pressure opens the highly compliant canals of young infants, resulting in fewer test ears with ear-canal collapse than the TPP condition. This is an inherent advantage of down-swept compared to up-swept tympanometric testing.

### 4.3 Did multivariate models improve test performance?

The best ambient predictors were absorbance and power level, combined into multivariate models with ART. Combining WAI variables into a multivariate predictor while constraining the number of input variables achieved generally equivalent performance on the evaluation samples. Ambient WAI variables combined with the ART test after outliers were excluded overall showed the best prediction (AUC= 0.78-0.81). The results obtained in the WBN group are similar to Sanford et al. (2009), who also studied WB multivariate models in well newborns.

The best tympanometric predictors were absorbance at TPP and admittances at positive tail pressures, especially when combined into multivariate models with ART. Generalization of the tympanometric multivariate predictor models for TEOAE and for Stage 2 screening was good, with AUC values in the validation samples equal to or exceeding the development samples in most cases. The models that included ART data consistently performed better than those without, thus inclusion of ART for newborn testing is helpful in detection of middle-ear problems. Because CHL changes the impedance characteristics of the middle ear, stimuli reaching the cochlea are reduced in level. However, ART may also be affected by sensory (cochlear) and neural loss, so comparison to WBT is needed to assess middle-ear pathology. ART is able to detect neural dis-synchrony for which OAE testing is insensitive, because neural dis-synchrony may be present in the absence of any outer hair cell dysfunction. In a previous report from this cohort (Hunter et al., 2016), the ART was elevated by 19 dB in ears that were found to have a CHL and by 15 dB for ears with a SNHL, each defined with diagnostic air and bone-conduction ABR. Comparisons of ART to wideband absorbance tympanometry and to OAE testing helps to distinguish between these hearing loss types within a diagnostic test battery. In contrast, WAI tests have the advantage of detecting conductive pathologies, which OAE tests alone cannot differentiate from cochlear pathologies. One caveat is that the ART test employed in this study was performed at TPP, for which the test instrument required a pressure pump. Whether an ambient ART response measured by a test instrument having no pressure pump would be as predictive has yet to be studied.

An important outcome was that the most accurate multivariate models in Table 3 were those based on WAI variables contained only at the four half-octave bin frequencies between 1 kHz and 2.8 kHz (as well as the ART). This is a specific frequency band in which conductive dysfunction was predicted in a large sample of newborns and is consistent with previous studies. Based on these results, it would be efficient to focus on this frequency range to test for conductive dysfunction in newborns.

In terms of predictive modeling, it was important to use two randomized groups in calculating test performance, modeled after Myers et al. (2018), refined by keeping both ears of an individual in one group due to their correlated results. Both AIC and BIC criteria were effective in limiting the number of inputs to each multivariate model so that the model’s predictive accuracy based on AUC was similar in the development and validation sub-groups. It was possible to achieve larger AUCs (exceeding 0.9) in the development sub-group with multivariate models including many more WAI variables. However, such high accuracy did not generalize to the validation sub-group, i.e., their resulting AUCs were much smaller than those of the best models.

### 4.4 Does detection of outliers improve prediction?

Based on distributions of each test variable, criterion values were established that indicated invalid tests consistent with acoustic signs of probe leaks or blockages. The ambient pressure test resulted in a larger number of outliers (19.6%) than the tympanometric test (13.5%). Exclusion of these outliers improved the AUCs for ambient tests in both the development and validation groups, indicating the validity of these exclusions for identifying inadequate probe fits. In the tympanometric tests, exclusion of outliers either did not change, or slightly decreased the AUCs, supporting our hypothesis that pressurization helps to ensure that an adequate probe fit has been obtained.

AlMakadma and Prieve (2021) reported that leaky probe fits contributed to increased absorbance and variability primarily in frequencies below 1 kHz, with minor differences in the region above 2 kHz. The best predictive models in the current and previous studies were for WAI variables in the frequency range from 1 to 2.8 kHz that are less impacted by leaky probe fits. However, the present results showed that this frequency range is impacted by blocked probes, thus outlier tests in this important frequency range are important to predict outer and middle-ear status in newborns. This is the first study to evaluate the application of probe-fit exclusion criteria on ambient or tympanometric predictive values of newborn screening tests, so further research is needed to validate these criteria for prediction models. This finding could improve hearing screening tests by measuring absorbance to test the fit of the probe tip and prevent false-positive referrals due to ear canal blockage.

### 4.5 Do the WB models perform as well in NICU infants?

In the WBN group, results were generally better than in the NICU infants, and exclusion of outliers improved prediction in the NICU group for the TEOAE test, but not as much for the Stage 2 test. When using the WB test battery in the NICU setting, it appears important to exclude outliers and include an ART, similar to the general pattern for the WBN. A caveat is that all models were developed in Group 1 with similar proportions of WBN and NICU ears as for Group 2 validation. The fact that the AUC2 was usually larger for WBN than all ears in the ambient WAI battery suggests that the models would have performed better on WBN ears if the ambient predictors had been developed only on WBN ears. The ART exam is important to achieve more accurate results in NICU ears on both the ambient and tympanometric test batteries. Conclusions regarding test performance in NICU ears are to be viewed with caution, because of the small number of refer ears (n=14) in the Stage 2 exam groups of the Group 2 sample, which declined to only 8 ears when outliers were excluded.

## 5 Strengths, Limitations, and Directions for Future Research

Strengths of the study include the size and diversity of the sample and inclusion of infants screened in the NICU, thus increasing generalizability and real-world application. This is the largest study published thus far of WAI for prediction of middle-ear problems associated with newborn hearing screening failure. It added WAI tests (pressurized WAI, including ART) not previously evaluated, and additional admittance variables beyond absorbance and phase, coupled with an outlier test to detect inadequate probe fit. The enrollment was designed to maximize the sample of refer ears so that robust power to predict fail results was possible. The large sample of newborns in both pass and refer groups enabled an adequate sample size to characterize outliers, while excluding them from further analysis, and to construct randomized samples for algorithm development and validation. This is the first study to assess prediction of wideband acoustic reflexes, to include infants screened in the NICU, and to assess predictive accuracy with and without exclusion of tests based on quality of the acoustic response.

### 5.1 Limitations

A limitation of this study is the time lag between the performance of NHS exams and performance of WAI tests. A possible contributor to lower AUCs in the current study compared to some previous ones (e.g., Hunter et al., 2010; Myers et al., 2018; Sanford et al., 2009) is that the WAI test was performed separately from the NHS exam by a different examiner using a different test instrument and probe, thus the middle-ear condition could change during the time between tests.

The examiner in this study for the WBN was a trained research assistant who was not an audiologist and did not perform the NHS exam. This approach was used to reduce bias with the same examiner performing the NHS exam and the WAI test, and due to practical limitations for a large-scale study in which the health mission places higher priority on completing the NHS exam and allowing the infant bonding time with the parents, rather than completing study procedures. These limitations represent real-world conditions, since it is not practical to have an audiologist performing screening and study tests simultaneously in newborn screening programs.

Another limitation of this and other NHS studies is the lack of a gold standard for conductive dysfunction at the time of NHS. The cause of conductive dysfunction could include outer-ear debris, ear-canal collapse, middle-ear fluid, mesenchyme, or abnormal middle-ear pressure. Without radiographic or surgical confirmation, which was not possible for practical and ethical reasons, conductive dysfunction was defined based on a standard NHS exam with TEOAE and/or automated ABR.

### 5.2 Future Research Recommendations

Questions for future research might include the following. 1) Study whether real-time information from the WAI test can help to ensure adequate probe fit for OAE testing. 2) Assess whether a combination of WAI (with ART) tests followed by the NHS exam using the same probe improves interpretation of the results of the NHS exam. 3) Investigate whether WAI (with ART) testing is useful in diagnostic follow-up protocols following the initial NHS exam

## 6 Conclusions

Overall, pressurized WAI and ART testing is helpful for interpreting results from NHS exams in newborns. It assesses the risk for conductive impairment, whether transient at the time of infant testing or longer lasting. Tympanometric tests are more accurate at predicting NHS exam status when data at the positive tail pressure is used rather than at TPP. This may be specific to newborns and young infants, as positive air pressure opens the highly compliant ear canals of young infants. This test combination could prove useful in screening populations at elevated risk for neural and conductive loss, such as infants cared for in NICUs. The combination could also prove to be an excellent second stage screening in well-baby nurseries, as the risk for conductive versus sensorineural hearing loss would be clearer. Using an ambient WB test in combination with OAE screening could provide valuable information about ear-canal and signal transmission integrity that would improve performance of the OAE screening.

## Data Availability

All data produced are available online at https://data.mendeley.com/datasets/3zgxr6d4gy/1

## Appendix A. Statistical results for ambient test responses

The logits of all generalized linear regression models are expressed using Wilkinson notation. Each model listed was significant via a Chi^2 statistic versus a constant model. See Notes to Table 4 for further information on the WB variable names used in the logits.

### A.1 Statistical results for NHS exam type: TEOAE

#### A.1.1 All ears

##### A.1.1.1 Model input with no Reflex test

*BIC and AIC model criteria results identical*

Generalized linear regression model: logit(NRefer) ∼ Aa20*WLa20

Estimated Coefficients:

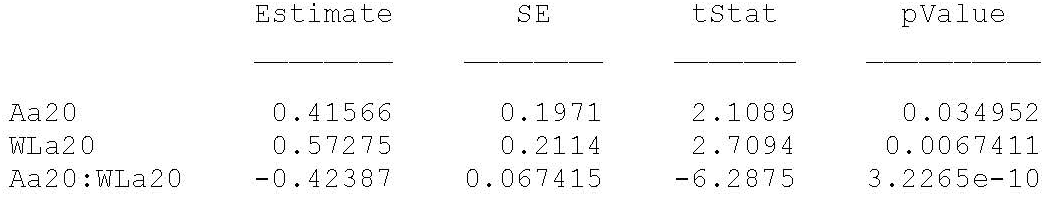

874 observations, 871 error degrees of freedom

##### A.1.1.2 Model input with Reflex test

*AIC model criterion*

Generalized linear regression model:

logit(Nrefer) ∼ Aa20 + ART + Aa14:Aa20 + Aa14:WLa20 + Aa20:WLa20 Estimated Coefficients:

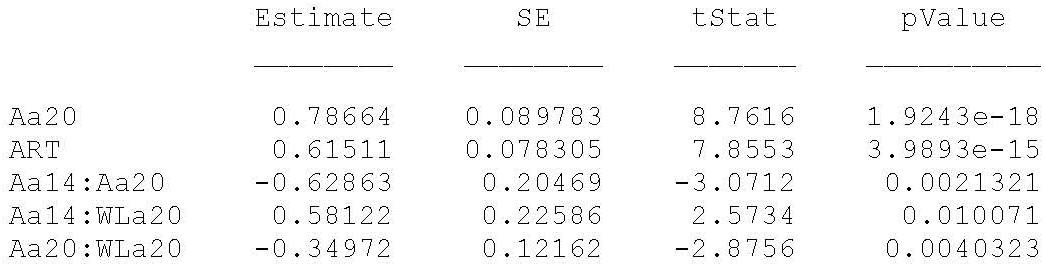

874 observations, 869 error degrees of freedom

#### A.1.2 Ears excluding outliers

##### A.1.2.1 Model input with no Reflex test

#### AIC model criterion

Generalized linear regression model:

logit(NRefer) ∼ Aa14 + WLa20 + Aa14:Aa20 + Aa14:WLa28 + Aa20:WLa28 + WLa20:WLa28

Estimated Coefficients:

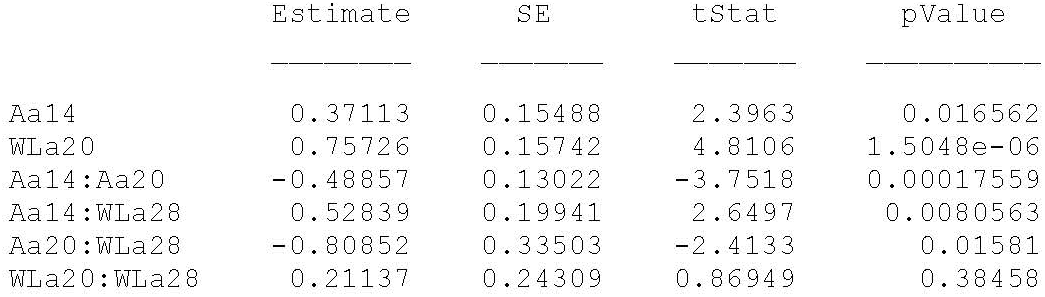

713 observations, 707 error degrees of freedom

##### A.1.2.2 Model input with Reflex test

*AIC model criterion*

Generalized linear regression model:

logit(Nrefer) ∼ 1 + ART + Aa10:Aa14 + Aa14:Aa20 + Aa20:Aa28 + Aa10*WLa20 + Aa14:WLa20 + Aa14:WLa28 + Aa20:WLa28

Estimated Coefficients:

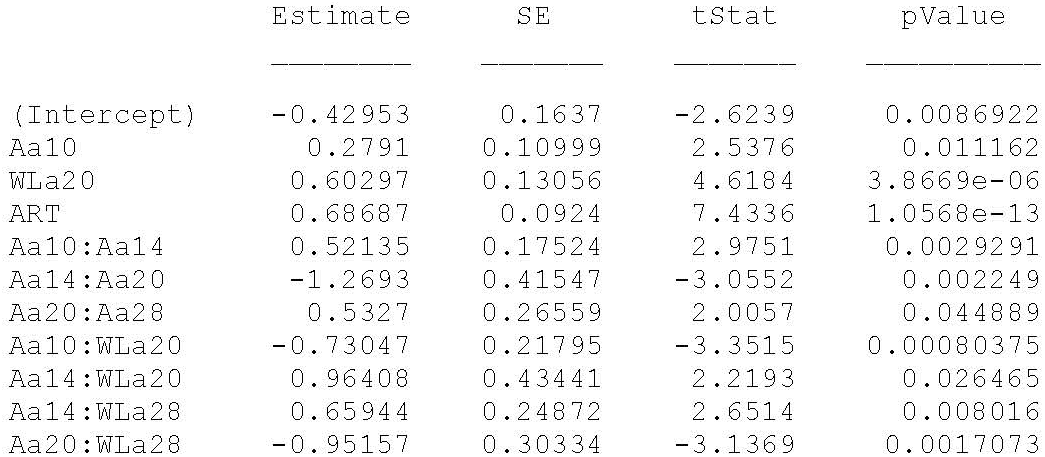

713 observations, 702 error degrees of freedom

### A.2 Statistical results for NHS exam type: 2stage

#### A.2.1 All ears

##### A.2.1.1 Model input with no Reflex test

*AIC and BIC model criteria lead to same model*

Principal Component Analysis Input WB variables to PCA:

Aa14, Aa20, Aa28, WLa14, WLa20

Corresponding output variables from PCA: PCa6, PCa7, PCa8, PCb6, PCb7

Orthonormal coefficient matrix

(input WB vars. in rows, output PCs in columns):

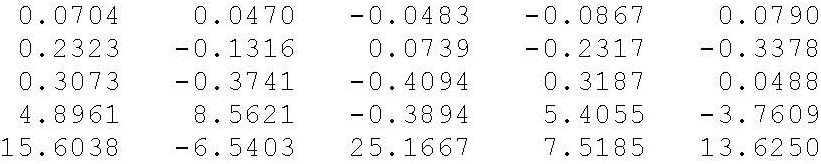

Variance explained by each principal component: 83.8692

10.0066

3.4280

2.2030

0.4932

Generalized linear regression model: logit(NRefer) ∼ 1 + PCa6 + PCb6 + PCa7:PCa8 Distribution = Binomial

Estimated Coefficients:

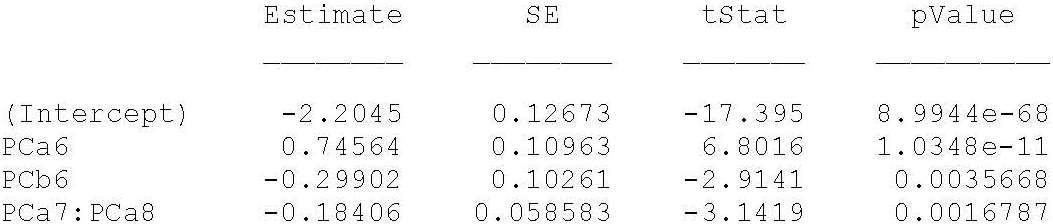

874 observations, 870 error degrees of freedom

##### A.2.1.2 Model input with Reflex test

*AIC model criterion*

Principal Component Analysis

Input WB variables to PCA:

Aa14, Aa20, Aa28, WLa14, WLa20

Corresponding output variables from PCA: PCa6, PCa7, PCa8, PCb6, PCb7

Orthonormal coefficient matrix

(input WB vars. in rows, output PCs in columns):

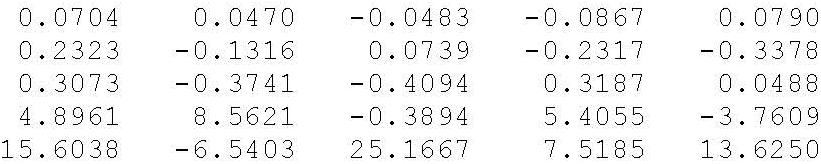

Variance explained by each principal component: 83.8692

10.0066

3.4280

2.2030

0.4932

Generalized linear regression model: logit(Nrefer) ∼ 1 + PCa7:PCa8 + PCa6*ART

Estimated Coefficients:

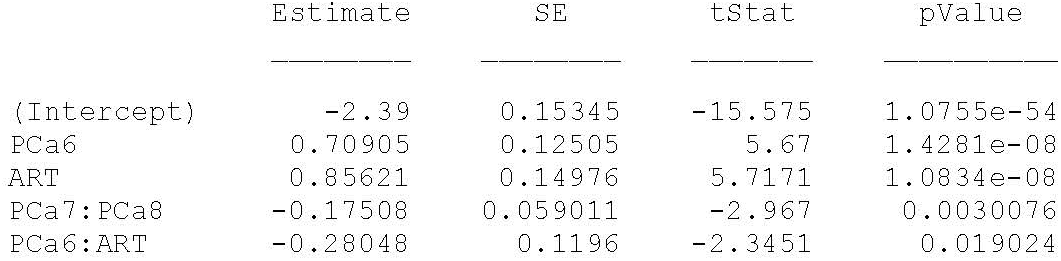

874 observations, 869 error degrees of freedom

#### A.2.2 Ears excluding outliers

##### A.2.2.1 Model input with no Reflex test

*AIC and BIC model criteria lead to same model*

Input variables to PCA: Aa14, Aa20, WLa14, WLa20

Output PC variables from PCA: PCa6, PCa7, PCb6, PCb7

Orthonormal coefficient matrix

(input vars. in rows, output vars. in columns):

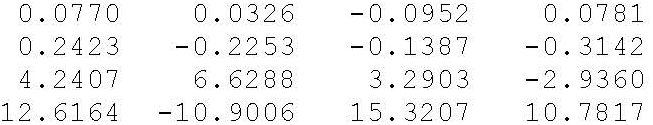

Variance explained by each principal component: 85.9138

10.2104

3.2508

0.6250

Generalized linear regression model: logit(NRefer) ∼ 1 + PCa6 + PCa6:PCb7 Distribution = Binomial

Estimated Coefficients:

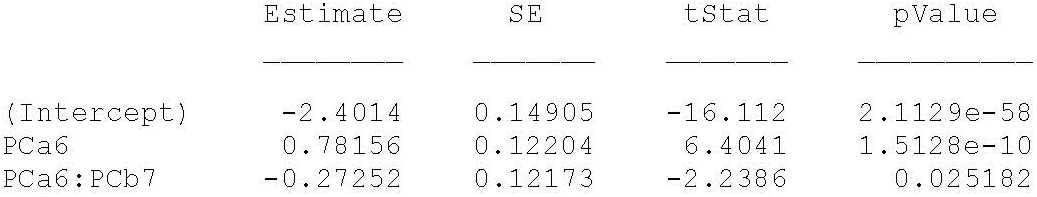

713 observations, 710 error degrees of freedom

##### A.2.2.2 Model input with Reflex test

*AIC model criterion*

Input variables to PCA: Aa14, Aa20, WLa20

Output variables from PCA: PCa6, PCa6, PCb7

Orthonormal coefficient matrix

(input vars. in rows, output vars. in columns):

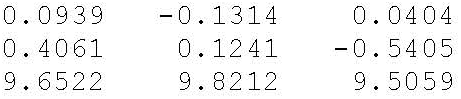

Variance explained by each principal component:

91.5758

6.5521

1.8720

Generalized linear regression model: logit(Nrefer) ∼ 1 + PCa6 + ART

Estimated Coefficients:

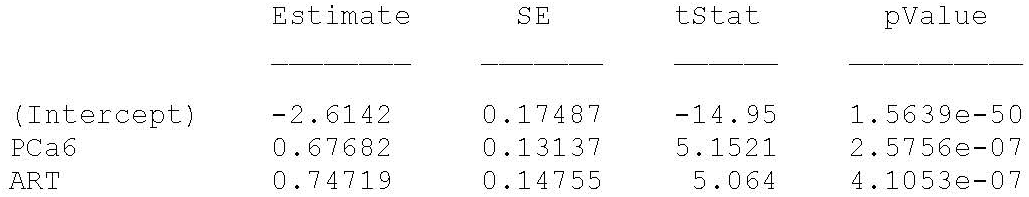

713 observations, 710 error degrees of freedom

## Appendix B. Statistical results for tympanometric test responses

### B.1 Statistical results for NHS exam type: TEOAE

#### B.1.1 All ears

##### B.1.1.1 Model input with no Reflex test

*AIC model criterion*

Generalized linear regression model:

logit(NRefer) ∼ YIpt10 + YIpt14 + YIpt28 + Atpp20 + YRpt28:YIpt10 + YRpt20:YIpt14 + YRpt28:YIpt14 + YRpt20:YIpt28 + YRpt20:Atpp14 + YRpt28:Atpp20

Estimated Coefficients:

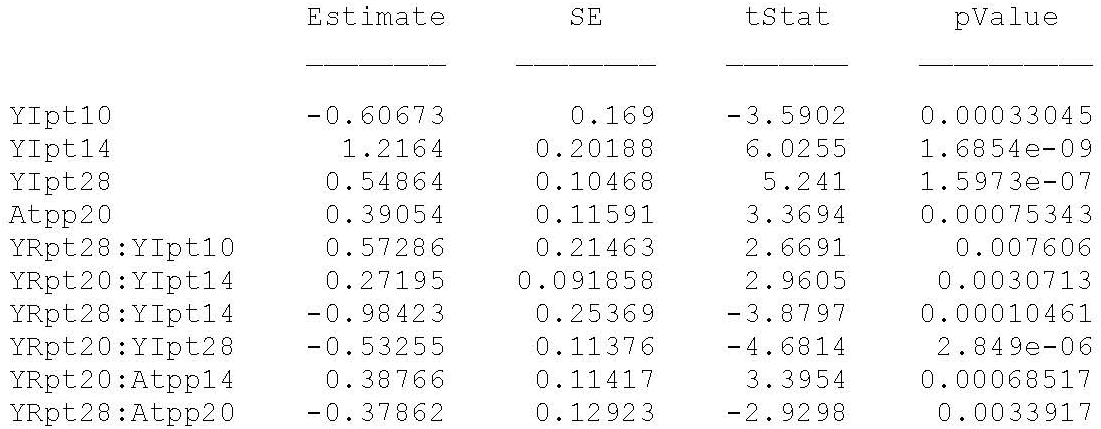

874 observations, 864 error degrees of freedom

##### B.1.1.2 Model input with Reflex test

*AIC model criterion*

Generalized linear regression model:

logit(Nrefer) ∼ 1 + YRpt20:YRpt28 + YRpt28:YIpt14 + YRpt20:YMpt20 + YIpt14:YMpt20 + YRpt28:ART + YIpt14:ART + YMpt20*ART + Atpp20:ART

Estimated Coefficients:

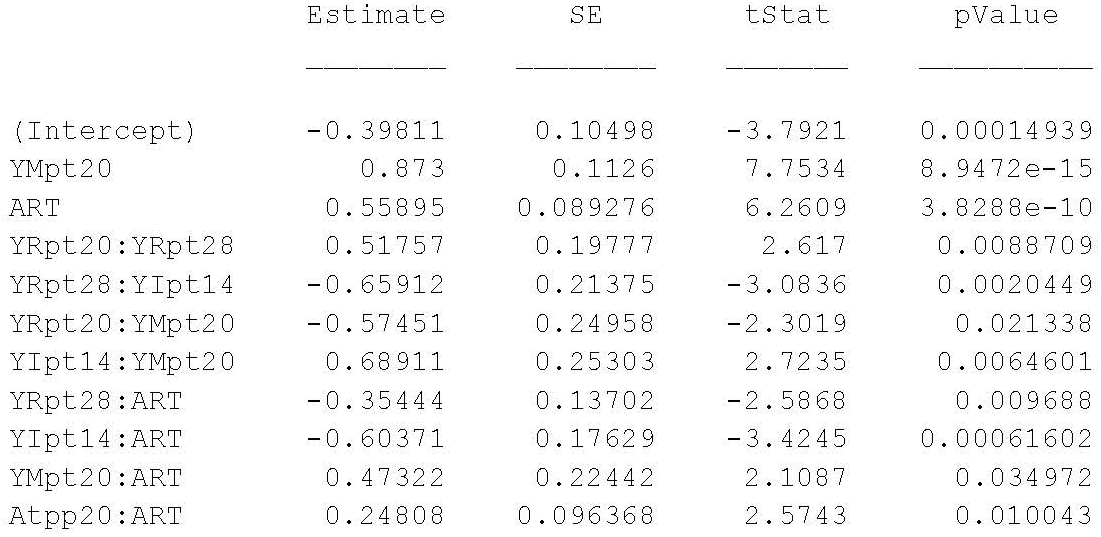

874 observations, 863 error degrees of freedom

#### B.1.2 Ears excluding outliers

##### B.1.2.1 Model input with no Reflex test

*AIC model criterion*

Generalized linear regression model:

logit(NRefer) ∼ 1 + YIpt14 + YRpt28:YIpt14 + YRpt28:YIpt28 + YRpt20:YMpt14 + YIpt28*YMpt20 + YMpt14*YMpt20

Estimated Coefficients:

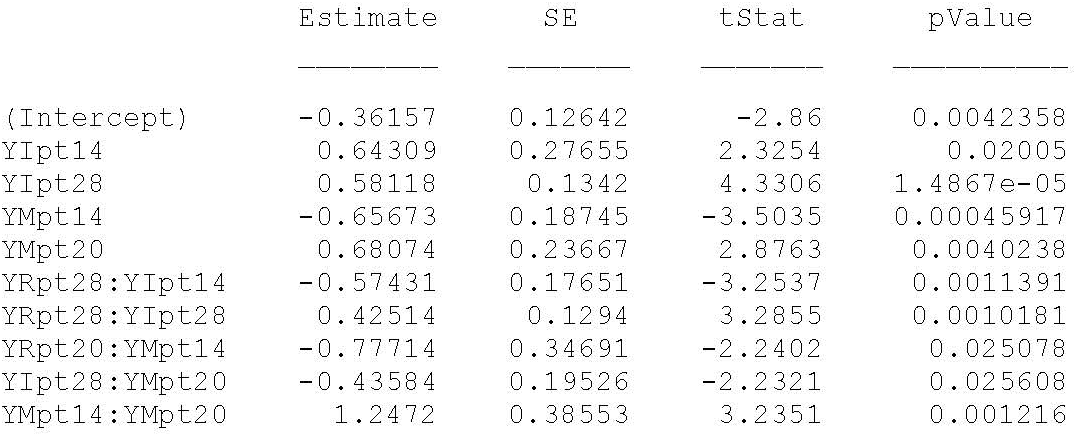

759 observations, 749 error degrees of freedom

### B.2. Ears excluding outliers

#### B.2.1 All ears

##### B.2.1.1 Model input with Reflex test

*BIC model criterion*

Generalized linear regression model: logit(Nrefer) ∼ 1 + YMpt20 + YIpt14*ART

Estimated Coefficients:

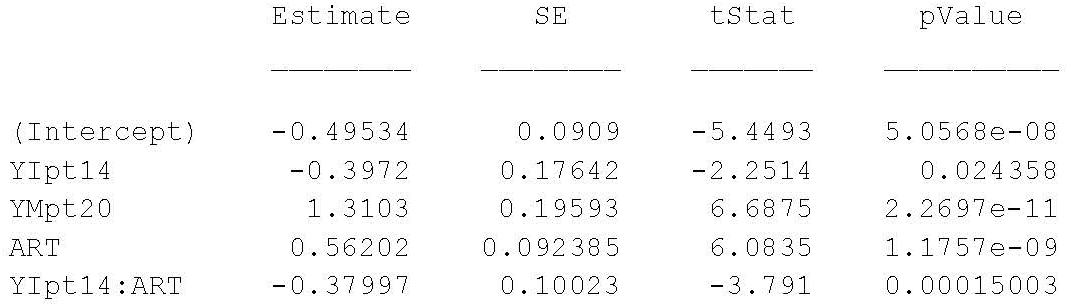

759 observations, 754 error degrees of freedom

### B.2 Statistical results for NHS exam type: 2stage

#### B.2.1 All ears

##### B.2.1.1 Model input with no Reflex test

*AIC model criterion*

Generalized linear regression model:

logit(NRefer) ∼ 1 + YMpt20 + YRpt28:YIpt14 + YIpt07:YIpt14 +

YRpt20:YMpt14 + YIpt14:YMpt14 + YRpt28:YMpt20 + YIpt07:YMpt20 +

YIpt14:YMpt20

Estimated Coefficients:

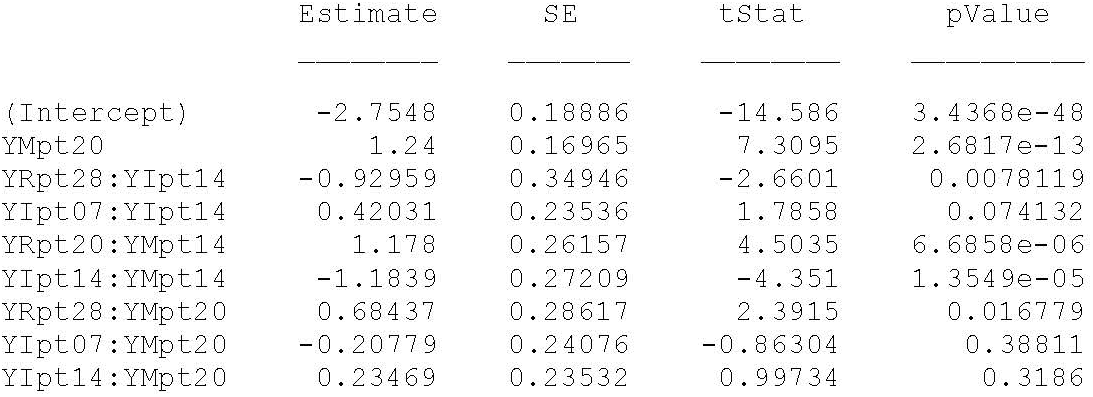

874 observations, 865 error degrees of freedom

##### B.2.1.2 Model input with Reflex test

*AIC model criterion*

Principal Component Analysis of WB variables

Input WB variables to PCA:

YRpt20, YRpt28, YIpt07, YIpt10, YIpt14, Atpp14, Atpp20

Output PCs from PCA:

PCa7, PCa8, PCb4, PCb5, PCb6, PCc6, PCc7

Orthonormal coefficient matrix

(input WB vars. in rows, output PCs in columns):

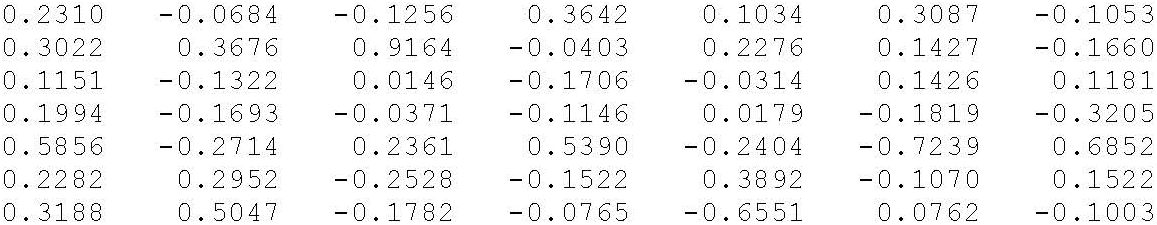

Variance explained by each principal component:

58.4848

17.0750

10.4683

7.8375

3.0081

1.7831

1.3433

Generalized linear regression model:

logit(Nrefer) ∼ 1 + PCb5 + PCa7:PCb4 + PCa8:PCb5 + PCb4:PCb6 +

PCb5:PCb6 + PCa7:PCc6 + PCb4:PCc6 + PCa7*ART + PCc7*ART

Estimated Coefficients:

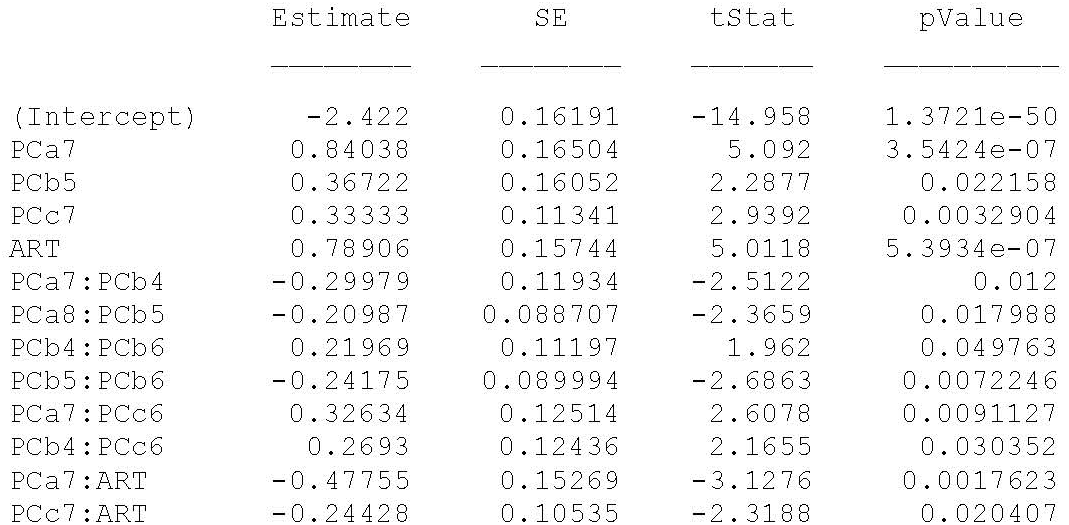

874 observations, 861 error degrees of freedom

#### B.2.2 Ears excluding outliers

##### B.2.2.1 Model input with no Reflex test

*BIC model criterion*

Generalized linear regression model:

logit(NRefer) ∼ 1 + YRpt20 + YRpt28

Estimated Coefficients:

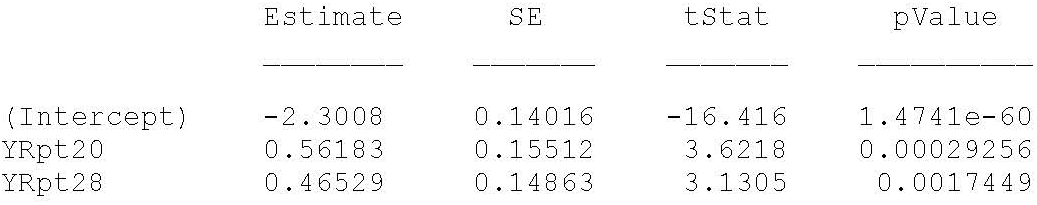

759 observations, 756 error degrees of freedom

##### B.2.2.2 Model input with Reflex test

AIC model criterion

Generalized linear regression model:

logit(Nrefer) ∼ 1 + YRpt28 + YMpt20 + YRpt20:YRpt28 + YRpt20:YMpt20 +

YIpt14:YMpt20 + YRpt20:YMpt28 + YMpt28*ART

Estimated Coefficients:

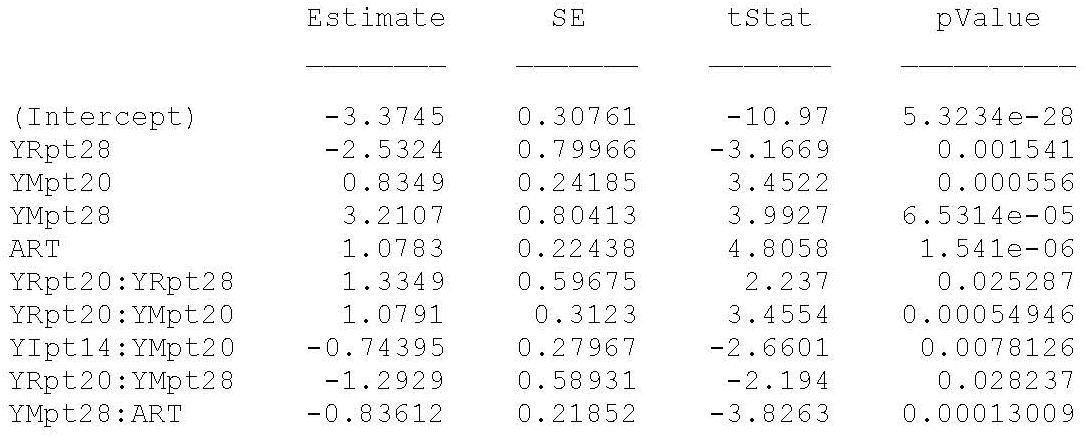

759 observations, 749 error degrees of freedom

## List of Abbreviations and Term definitions

a: Suffix appended to indicate ambient-pressure variables
Aa: Absorbance at ambient pressure
ABR: Auditory Brainstem Response
AIC: Akaike Information Criterion
Apt: Absorbance at positive pressure tympanometry tail
ART: Acoustic Reflex Threshold
ASR: Acoustic Stapedial-muscle Reflex
Atpp: Absorbance at Tympanometric Peak Pressure
AUC: Area Under the Receiver Operating Characteristic Curve
BBN: Broad Band Noise
BIC: Bayesian Information Criterion
D: Group delay (phase expressed in time units across frequencies)
DPOAE: Distortion Product Otoacoustic Emissions
GLM: General Linear Model
IQR: Inter-Quartile Range
NHS: Newborn Hearing Screening
NICU: Newborn Intensive Care Unit
NN: Number Normal
nt: Suffix appended to indicate negative-tail tympanometric variables
PCA: Principal Component Analysis
Pnt: Pressure at negative pressure tympanometry tail
Ppt: Pressure at positive pressure tympanometry tail
pt: Suffix appended to indicate positive-tail tympanometric variables
ROC: Receiver Operating Characteristic
SNHL: Sensorineural Hearing Loss
SNR: Signal-to-Noise Ratio
SPL: Sound Pressure Level
TEOAE: Transient Evoked Otoacoustic Emissions
TPP: Tympanometric Peak Pressure
WAI: Wideband Acoustic Immittance
WBT: Wideband Tympanometry
WBN: Well-baby nursery
WL: Absorbed Power Level
YI: Susceptance or imaginary part of admittance
YM: Admittance Magnitude
YP: Admittance Phase
YR: Conductance or real part of admittance

